# Transcriptome Signatures for Cognitive Resilience Among Individuals with Pathologically Confirmed Alzheimer Disease

**DOI:** 10.1101/2024.11.12.24317218

**Authors:** Donghe Li, Xudong Han, Lindsay A. Farrer, Thor D. Stein, Gyungah R. Jun

## Abstract

**INTRODUCTION:** Limited success to date in development of drugs that target hallmark Alzheimer disease (AD) proteins as a means to slow AD-related cognitive decline has sparked interest in approaches focused on cognitive resilience. We sought to identify transcriptome signatures among brain donors with neuropathologically confirmed AD that distinguish those with cognitive impairment from those that were cognitively intact.

**METHODS:** We compared gene expression patterns in brain tissue from donors in four cohorts who were cognitively and pathologically normal (controls), met clinical and pathological criteria for AD (SymAD), or were cognitively normal prior to death despite pathological evidence of AD (cognitively resilient or AsymAD). Differentially expressed genes (DEGs) at the transcriptome-wide significance (TWS) level (P<10^-6^) in the total sample and nominally significant (P<0.05) in at least two datasets were further evaluated in analyses testing association of gene expression with co-calibrated and harmonized cognitive domain scores and AD-related neuropathological traits.

**RESULTS:** We identified 52 TWS DEGs, including 14 that surpassed a significance threshold of P<5×10^−8^. The three most significant DEGs, *ADAMTS2* (Log2 fold change [Log2FC]=0.46, P=2.94×10^−14^), *S100A4* (Log2FC=0.61, P=3.98×10^−11^) and *NRIP2* (Log2FC=0.32, P=9.52×10^−11^) were up-regulated in SymAD compared to AsymAD brains. *ADAMTS2* and *SLC6A9* were also significantly and nominally differentially expressed between AsymAD cases and controls (FDR P=0.45 and FDR P=0.57, respectively). Significant associations (P<0.0038) were identified for executive function with expression of *ADAMTS2* (P=4.15×10^−8^) and *ARSG* (P=1.09×10^−3^), and for memory with *PRELP* (P=3.92×10^−5^) and *EMP3* (P=7.75×10^−4^), and for language with *SLC38A2* (P=6.76×10^−5^) and *SLC6A9* (P=2.13 ×10^−3^). Expression of *ARSG* and *FHIP1B* were associated with measures of Tau pathology (AT8: P=1.5×10^−3^, and pTau181: P=3.64×10^−3^, respectively), and *SLC6A9* expression was associated with multiple pTau isoforms including pTau181 (P=1.5×10^−3^) and pTau396 (P=2.05×10^−3^). *PRELP* expression was associated with synaptic density *(PSD.95: P=6.18*×10^−6^). DEGs were significantly enriched in pathways involving E2F targets, cholesterol homeostasis, and oxidative phosphorylation.

**CONCLUSION:** We identified multiple DEGs that differentiate neuropathologically confirmed AD cases with and without cognitive impairment prior to death. Expression of several of these genes was also associated with measures of cognitive performance and AD-related neuropathological traits, thus providing important insights into cognitive resilience mechanisms and strategies for delaying clinical symptoms of AD.

## Background

The growth in life expectancy has resulted in a rapidly aging population, which is accompanied by an increase in age-related diseases such as Alzheimer disease (AD) [1]. AD, the most common cause of dementia globally, is characterized by progressive accumulation of extracellular amyloid plaques with amyloid-β (Aβ) protein and intraneuronal neurofibrillary tangles due to intracellular hyperphosphorylated tau filaments in the brain [2–4], which cause gradual neuronal degeneration and cognitive decline [2]. In addition, AD is associated with failure in a variety of biological process, including synaptic transmission, glial-mediated immunity, and mitochondrial metabolism [5–9].

Previous studies revealed that clinical indicators of cognitive decline appear after several years of neuropathological changes [10, 11]. Despite the presence of a large number of Aβ plaque aggregates and tau neurofibrillary tangles upon postmortem neuropathological examination consistent with an AD diagnosis, some individuals who meet pathological criteria for AD remain cognitively normal, and these individuals are often referred to as asymptomatic AD [11, 12]. Understanding the mechanisms underlying preserved cognitive health among AsymAD individuals (i.e., cognitive resilience) may provide insight as well as potential novel therapeutic targets for inhibiting symptomatic conversion.

To identify molecular patterns that distinguish symptomatic and asymptomatic individuals with neuropathologically confirmed AD, we conducted a transcriptome-wide differential gene expression analysis and evaluated association of the top-ranked differentially expressed genes (DEGs) with harmonized performance measures for several cognitive domains and with AD-related neuropathological traits in autopsied brains. Findings from these analyses informed subsequent examination of biological pathways that may be related to mechanisms involved in cognitive resilience.

## Methods

### Study participants and diagnostic criteria

Clinical and neuropathological data for 944 brain donors were obtained from the Framingham Heart Study (FHS, n=127), Boston University Alzheimer’s Disease Research Center (BUADRC, n=85), Religious Orders Study and Rush Memory and Aging Project (ROSMAP, n=567), and the Mount Sinai Brain Bank (MSSB, n=165) [13–17]. Neuropathological assessments of FHS and BUADRC subjects were carried out in accordance with protocols and guidelines developed and harmonized across studies at the Department of Veterans Affairs-Boston University brain bank [18–23]. AD cases were classified as symptomatic AD (SymAD) and cognitively resilient (AsymAD) based on cognitive assessment prior to death and neuropathological diagnosis. A pathological diagnosis of AD was established based on intermediate to high likelihood of AD using the NIA-Reagan criteria, and a clinical AD diagnosis was determined by Clinical Dementia Rating (CDR) score (greater than 0.5) or Clinical cognitive diagnosis scores (4 and 5). A control group included participants who were neither neuropathologically nor clinical diagnosed with AD. Using this classification system, the total sample included 385 SymAD cases, 208 AsymAD cases and 351 controls, all of whom were of European ancestry. Brain donor characteristics in each dataset are shown in **Table 1**.

**Table 1.** Characteristics of participants by dataset and diagnostic group.

| Dataset<br>Variables | ROSMAP (N=567) |  |  | MSBB (N=165) |  |  | FHS (N=127) |  |  | BUADRC (N=85) |  |  |
| --- | --- | --- | --- | --- | --- | --- | --- | --- | --- | --- | --- | --- |
|  | SymAD | AsymAD | Control | SymAD | AsymAD | Control | SymAD | AsymAD | Control | SymAD | AsymAD | Control |
| **N** | 200 | 172 | 195 | 113 | 4 | 48 | 42 | 12 | 73 | 30 | 20 | 35 |
| **Number males/**<br>**Number females** | 59/141 | 61/111 | 76/119 | 40/73 | 2/2 | 18/30 | 15/27 | 5/7 | 37/36 | 16/14 | 9/11 | 20/15 |
| **Mean age at**<br>**death (SD)** | 91.0<br>(5.9) | 88.8<br>(5.7) | 86.2<br>(6.9) | 84.2<br>(6.9) | 89.5<br>(1.0) | 82.2<br>(8.8) | 90.4<br>(7.2) | 88.4<br>(9.6) | 85.6<br>(10.1) | 82.0<br>(9.9) | 82.1<br>(8.1) | 84.9<br>(8.3) |
| **Mean Braak**<br>**Stage (SD)** | 4.36<br>(0.85) | 3.89<br>(0.76) | 2.46<br>(1.11) | 5.30<br>(1.03) | 4.25<br>(0.96) | 1.90<br>(1.13) | 5.14<br>(0.90) | 4.25<br>(0.97) | 1.95<br>(0.85) | 5.40<br>(0.77) | 5.05<br>(0.89) | 2.40<br>(1.19) |
| **Mean CERAD**<br>**score (SD)** | 2.59<br>(0.49) | 2.35<br>(0.49) | 0.41<br>(0.63) | 2.76<br>(0.47) | 2.00<br>(0.82) | 0.33<br>(0.60) | 2.62<br>(0.58) | 2.33<br>(0.78) | 0.60<br>(0.74) | 2.47<br>(0.70) | 2.07<br>(0.83) | 0.52<br>(0.81) |
| **Number APOE**<br>**ε4 carriers (%)** | 81<br>(41) | 45<br>(26) | 20<br>(10) | 47<br>(42) | 2<br>(50) | 11<br>(23) | 17<br>(40) | 5<br>(42) | 11<br>(15) | 23<br>(77) | 8<br>(40) | 9<br>(26) |

SD: Standard deviation.

### RNA-seq data acquisition and processing

RNA-sequencing (RNA-seq) data for all subjects were derived from dorsolateral prefrontal cortex (DLPFC) tissue. Data for ROSMAP and MSBB subjects were obtained from the CommonMind Consortium portal (http://www.synapse.org). Details about procedures for preprocessing and quality control of the RNA-seq data and normalization of the gene expression data for ROSMAP, MSBB and FHS subjects are reported elsewhere [16, 17, 24]. BUADRC RNA-seq data were processed using the same QC procedure [24]. Informed consent for all participants was obtained under protocols approved by the institutional review boards from Boston University, the VA Bedford Healthcare System (Bedford, MA), Rush University Medical Center, and the Mount Sinai and JJ Peters VA Medical Center.

### Calculation of cognitive domain scores

Co-calibrated harmonized scores for memory, language and executive function domains were calculated from non-identical cognitive test batteries administered to ROSMAP, FHS, and BUADRC subjects, and these scores were standardized on a unified scale as previously described [25–28]. Briefly, this process involved the meticulous assignment of neuropsychological test items to their respective domains by a panel of experts [25], followed by confirmatory factor analysis utilizing Mplus [29] software for calibration purposes. Subsequently, scores for each domain were computed independently using data from the most recent evaluation, with test items common across multiple cohorts serving as anchor points for calibration. The parameters of these anchor items were harmonized across studies to ensure the comparability of scores. Additionally, cognitive scores with a standard error exceeding 0.6 or derived solely from the mini-mental state examination (MMSE) were excluded from the analysis. The higher values of the scores indicate better performance of cognitive function. A summary of cognitive domain scores is shown in **Supplementary Table 1**.

### Brain protein measurement procedures

Several proteins were measured in fixed in periodate-lysine-paraformaldehyde brain tissue from FHS and BUADRC donors. Tissue blocks were paraffin-embedded and sections were cut at 10 µm for immunohistochemistry [19, 20]. Antigen retrieval for β-amyloid (Aβ) was performed with formic acid treatment for two minutes. Sections were incubated overnight at 4°C with antibodies to phosphorylated paired helical filament (PHF) tau (AT8; Pierce Endogen, Rockford IL; 1:2000), ionized calcium binding adaptor molecule 1 (Iba1) (Wako, 1:500). Quantification of AT8 and Iba1 cellular density was performed using previously described methods [30] with ImageScope software (Leica Biosystems) [31]. Levels of phosphorylated tau at 181 (pTau181), 202 (pTau202), 231 (pTau231) and 396 (pTau396), postsynaptic density protein 95 (PSD-95), Alpha-synuclein (aSyn), β-amyloid 42 and 40 (Aβ42 and Aβ40) from the DLPFC (Brodmann area 8/9) were measured by immunoassays using procedures reported elsewhere [32, 33]. Summarized information for the neuropathological traits and proteins assayed in brain is shown in **Supplementary Table 2**.

### Differential gene expression analysis

An adjustment for removing unwanted variations (RUVs) in gene expression [34] to control for batch, library preparation and other nuisance effects was derived using DEseq2 which applies a median of ratios normalization method [35]. Using the normalized expression counts from this process, differential expression (DE) of 16,842 protein coding genes between SymAD and AsymAD cases were evaluated separately in each of the four datasets by a regression model that included covariates for sex and age at death. DE results from the four datasets were combined by meta-analysis using a procedure incorporated in the METAL program [36] which assigns log2 fold change (log2FC) as the effect size and the standard error of log2FC as the standard error. We used “SCHEME STDERR” commands to weight effect size estimates using the inverse of the corresponding standard errors. Differentially expressed genes (DEGs) at the transcriptome-wide significance (TWS) level (P<10^−6^) in meta-analysis and those that were nominally significant (P<0.05) in at least two datasets were included in subsequent analyses. Gene enrichment analysis seeded with DEGs that were nominally significant (P<0.05) in the total sample was conducted using the Enrichr [37] package in R and pathway gene sets in the Molecular Signatures 2020 [38, 39] and KEGG Human 2021[40] databases. Pathway enrichment P-values were adjusted for the FDR with an adjusted P-value threshold of 0.05.

### Association of cognitive domain scores and AD-related neuropathological traits and proteins with DEGs

Expression of significant DEGs was tested for association with domain scores in memory, language, and executive function in the ROSMAP, FHS, and BUADRC datasets, and the results from each dataset were combined by meta-analysis. Models testing the association of gene expression levels with cognitive domain scores included covariates for sex, age at exam, education and RUVs. Association of gene expression with measures of tau, β-amyloid, synaptic density and neuroinflammation that were rank-transformed after adjusting for sex and age at death as previously described [41] was evaluated in FHS and BUADRC subjects, both in total and subgroups of SymAD and AsymAD using a linear regression model including a covariate for RUVs.

The study design and sequence of analyses are illustrated in **Figure 1**.

**Figure 1.**
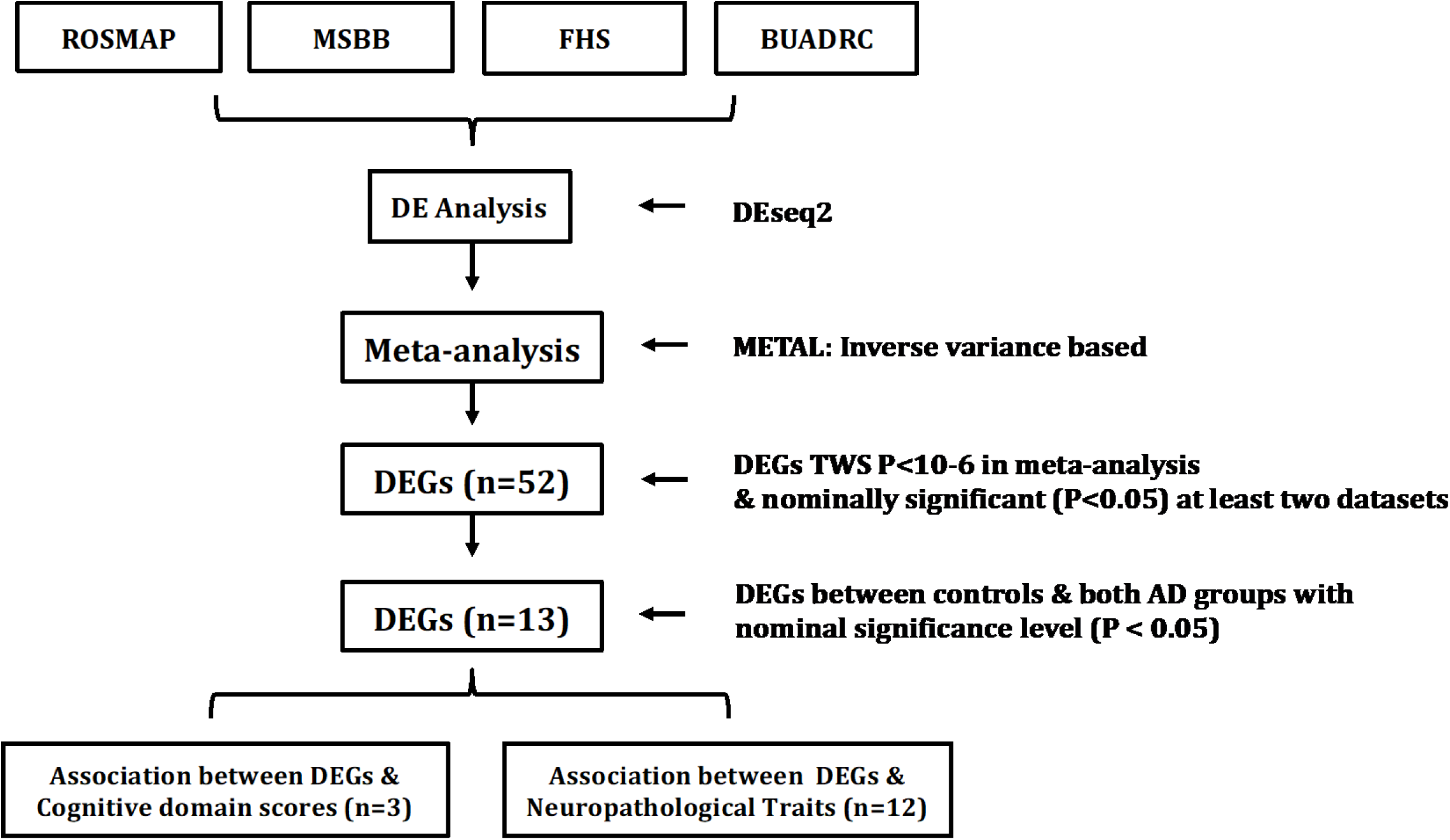
Study design and sequence of analyses. DE: differential expression. DEG: differentially expressed gene.

## Results

### Genes differentially expressed between SymAD and AsymAD cases

DE analysis revealed 52 genes with P-values < 1×10^−6^ in meta-analysis and < 0.05 in at least two of the datasets (**Figures 2A and B**). Among these, 14 genes were significant at the genome-wide level (P < 5×10^−8^, **Supplementary Table 3**). The four most significant DEGs, *ADAMTS2* (Log2FC=0.46, P=2.94×10^−14^), *S100A4* (Log2FC=0.61, P=3.98×10^−11^), *NRIP2* (Log2FC=0.32, P=9.52×10^−11^) and *SCGN* (Log2FC=0.55, P=2.55×10^−10^), were at least nominally significant in all four datasets (**Supplementary Table 3**) and were up-regulated in SymAD compared to AsymAD brains. Among the other 10 genes in this group, 7 (*SLC38A2*, *CHGA*, *VAT1*, *PAFAH1B3*, *POLD1*, *ARG2*, *SLC6A9*) were up-regulated and three (*ALDH1A1*, *CEP83*, *IVD*) were down-regulated (**Figure 2B and Supplementary Table 3**). To test the robustness of these results, we also performed a sensitivity analysis for the 52 DEGs by adjusting for AD-related covariates including Braak stage, CERAD score, and *APOE ɛ*4 carrier status. These analyses did not reveal any substantive differences among the results (**Supplementary Table 4**). Gene enrichment analyses that were seeded with nominally significant DEGs identified pathways involving the E2F targets, cholesterol homeostasis, and oxidative phosphorylation (**Supplementary Figure 1**).

**Figure 2:**
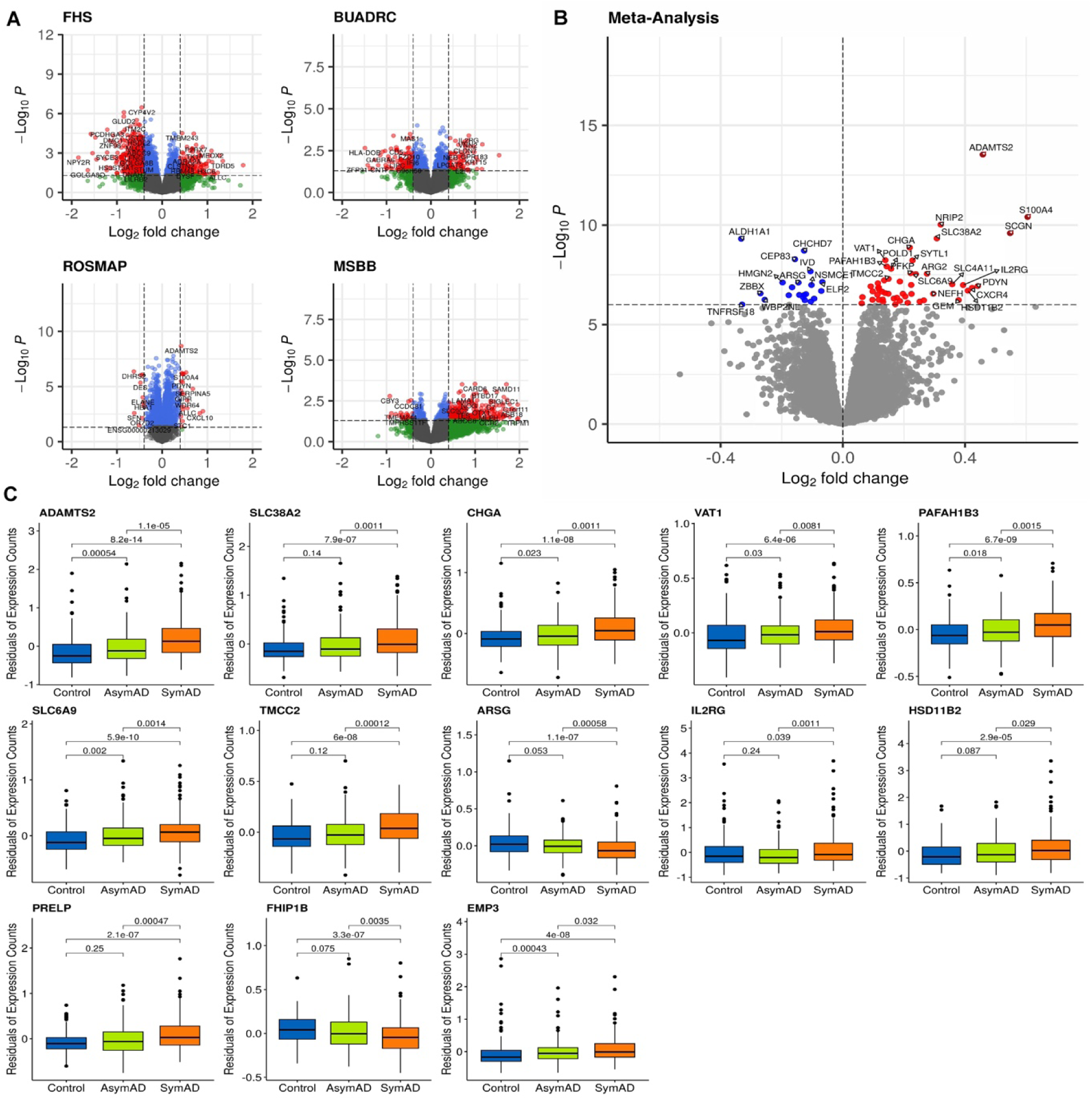
Differentially expressed genes identified between symptomatic and asymptomatic AD cases in **(A)** each individual dataset and (**B)** total sample. Red dots denote upregulated genes and blue dots represent downregulated genes at the transcriptome-side significance (TWS) level (P < 1.0×10^−6^). (**C)** Expression of TWS differentially expressed genes in controls, AsymAD cases and SymAD cases in the ROSMAP dataset which were also significantly differentially expressed between AsymAD cases and controls in the total sample.

Thirteen of the 52 transcriptome-wide significant DEGs between SymAD and AsymAD cases were also differentially expressed between controls and both AD groups (AsymAD and SymAD) at a nominal significance level (P<0.05; **Table 2**). Expression differences between AsymAD cases and controls remained significant after Bonferroni correction (P≤0.0038) for *ADAMTS2* (Log2FC=0.21, P=8.73×10^−4^) and *SLC6A9*(Log2FC=0.13, P =2.18×10^−3^). In this group of 13 DEGs, a nominally significant trend of increased expression in the order Control → AsymAD → SymAD was observed for *ADAMTS2, SLC38A2, CHGA, VAT1, PAFAH1B3, SLC6A9*, *HSD11B2,* and *EMP3*, and for *ARSG* and *FHP1B* in the reverse order in ROSMAP Study brain donor tissue (**Figure 2C**).

**Table 2:** Top-ranked differentially expressed genes between SymAD and AsymAD cases.

| Gene | SymAD vs AsymAD |  |  | AsymAD vs Control |  |  | SymAD vs Control |  |  |
| --- | --- | --- | --- | --- | --- | --- | --- | --- | --- |
|  | Log2FC | LfcSE | P-value | Log2FC | LfcSE | P-value | Log2FC | LfcSE | P-value |
| *ADAMTS2* | 0.458 | 0.060 | 2.94E-14 | 0.211 | 0.064 | 8.73E-04 | 0.580 | 0.053 | 4.18E-28 |
| *SLC6A9* | 0.239 | 0.043 | 2.93E-08 | 0.125 | 0.041 | 2.18E-03 | 0.333 | 0.038 | 3.94E-18 |
| *ARSG* | -0.146 | 0.027 | 7.80E-08 | -0.072 | 0.027 | 7.93E-03 | -0.186 | 0.023 | 6.70E-16 |
| *HSD11B2* | 0.409 | 0.079 | 1.98E-07 | 0.215 | 0.082 | 8.46E-03 | 0.499 | 0.068 | 1.47E-13 |
| *EMP3* | 0.212 | 0.043 | 8.06E-07 | 0.120 | 0.048 | 1.32E-02 | 0.294 | 0.042 | 3.64E-12 |
| *PAFAH1B3* | 0.144 | 0.025 | 1.20E-08 | 0.061 | 0.025 | 1.36E-02 | 0.201 | 0.022 | 1.10E-20 |
| *PRELP* | 0.225 | 0.044 | 2.90E-07 | 0.098 | 0.044 | 2.38E-02 | 0.284 | 0.038 | 9.71E-14 |
| *TMCC2* | 0.149 | 0.027 | 5.06E-08 | 0.061 | 0.027 | 2.60E-02 | 0.173 | 0.024 | 4.05E-13 |
| *VAT1* | 0.139 | 0.024 | 5.85E-09 | 0.046 | 0.021 | 3.05E-02 | 0.171 | 0.021 | 2.03E-16 |
| *FHIP1B* | -0.121 | 0.024 | 2.95E-07 | -0.049 | 0.023 | 3.33E-02 | -0.105 | 0.019 | 5.22E-08 |
| *SLC38A2* | 0.308 | 0.049 | 4.71E-10 | 0.094 | 0.047 | 4.35E-02 | 0.331 | 0.041 | 1.37E-15 |
| *CHGA* | 0.219 | 0.036 | 1.35E-09 | 0.066 | 0.033 | 4.70E-02 | 0.214 | 0.032 | 1.18E-11 |
| *IL2RG* | 0.394 | 0.074 | 1.04E-07 | -0.151 | 0.077 | 4.94E-02 | 0.177 | 0.069 | 9.80E-03 |

Genes were also differentially expressed between controls and both AD groups at a nominal significance level ( $P < 0.05$ ) and are listed in order of significance of comparison of AsymAD cases vs controls. Results for each comparison were derived from meta-analyses of four cohorts.
Log2FC: log2 fold change. LfcSE: standard error of log2 fold change.

### Association of genes differentially expressed between SymAD and AsymAD with cognitive performance and AD-related neuropathological traits

Among the 13 significant DEGs for SymAD versus AsymAD, executive function was significantly associated with expression of *ADAMTS2* (β=-0.0021, P=9.91×10^−9^) and *ARSG* (β=0.0033, P=1.09×10^−3^), memory function was significantly associated with expression of *PRELP* (β=-0.0004, P=3.92×10^−5^) and *EMP3* (β=-0.0015, P=7.75×10^−4^), and language performance was significantly associated with expression of *SLC38A2* (β=-0.0002, P=6.76×10^−5^) and *SLC6A9* (β=-0.0009, P=2.13×10^−3^). These associations were also significant in at least two datasets at a nominal significance level (P < 0.05; **Figure 3 and Supplementary Table 5**). Among AD-related neuropathological traits, AT8 was significantly associated with expression of *ARSG* (β=-0.011, P=1.58×10^−3^) in both the combined and separate FHS and BUADRC datasets (**Supplementary Table 6**). Expression of *SLC6A9* was significantly associated with multiple pTau isoform levels including pTau181 (β=0.0018, P=1.5×10^−3^), pTau396 (β=0.0018, P=2.05×10^−3^), and pTau231 (β=0.0017, P=6.04×10^−3^; **Supplementary Table 6**).

**Figure 3.**
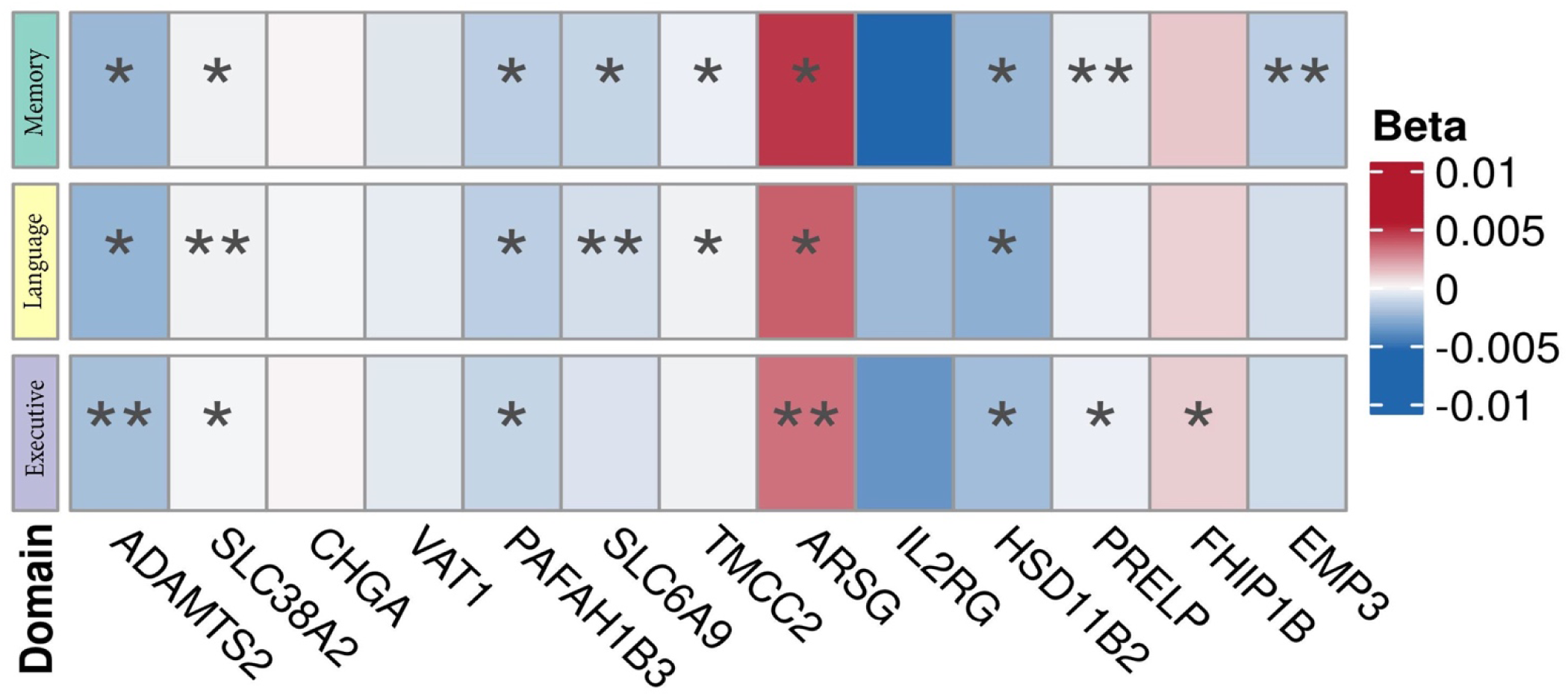
Association of expression of genes that are differentially expressed between SymAD and AsymAD cases with performance in memory, executive function and language in the ROSMAP, FHS, and BUADRC datasets. *: adjusted P < 0.05. **: nominally significant (P<0.05) in at least one additional dataset.

We further tested association of expression of *ADAMTS2* and *SLC6A9* with cognitive and brain tissue measures within 42 SymAD, 12 AsymAD, and 73 controls in the FHS dataset. *ADAMTS2* expression was significantly associated with pTau181, pTau231, Aβ42 and *ICAM1* levels in the SymAD (P < 0.05; **Supplementary Table 7**). *SLC6A9* expression was associated with pTau181 (β = 0.0021, P=2.01×10^−2^), pTau202 (β = 0.0026, P=1.07×10^−2^) and pTau396 (β = 0.0032, P=8.18×10^−4^) in SymAD cases. *SLC6A9* expression was also associated with CAA (β = 0.009, P=7.43×10^−3^) in AsymAD cases.

## Discussion

We identified 52 genes that were differentially expressed at a transcriptome-wide significance level in prefrontal cortex of pathologically confirmed AD cases of European ancestry who were cognitively impaired (SymAD) compared to those who were cognitively intact prior to death (AsymAD). Expression of 13 of these genes was also significantly different between AsymAD cases and persons without clinical or pathological signs of AD, and two genes showed a significant trend of increased expression according to disease severity (cognitively healthy → AsymAD → SymAD). These findings were not confounded by severity of Tau and Aβ pathology or *APOE* ɛ4 carrier status. Overall, this study identified novel genes associated with cognitive resilience to AD neuropathology.

The robustness and potential importance of *ADAMTS2*, the gene most significant differentially expressed between SymAD and AsymAD cases, is highlighted by a very recent DGE study in DLPFC tissue from a sample of 207 African American brain donors [42]. In that sample, *ADAMTS2* was the top differentially expressed gene between individuals with neuropathologically confirmed AD and those showing no evidence of AD. Another study, using a machine learning method with Boruta on ROSMAP transcriptome data, identified an increase in *ADAMTS2* levels associated with cognitive decline [43]. Although not previously implicated in AD genetic studies, several SNPs within *ADAMTS2* showed some evidence of association with AD risk (e.g., rs549839389, P=3.34×10^−4^; **Supplementary Table 8**) [44].

*ADAMTS2* encodes a member of the ADAMTS (a disintegrin and metalloproteinase with thrombospondin motifs 2) protein family. This particular enzyme, also referred to as procollagen I N-proteinase (PC I-NP) [45, 46], is involved in the processing and cleavage of procollagen molecules, which are precursors to collagen fibrils [47]. Collagen is an important structural component of the extracellular matrix in various tissues, including the brain [48]. Dysregulated extracellular matrices (ECM), which play a leading role in neuronal migration, axon guidance, and synaptic plasticity during development and in adulthood, could affect blood-brain-barrier integrity [43, 49–51]. We found that *ADAMTS2* expression was nominally associated with the level of pTau396 (P=0.036). Although direct links between *ADAMTS2* and pTau396 have not been reported, some studies showed that ECM proteins can modulate the aggregation and clearance of abnormally phosphorylated tau [52, 53]. *ADAMTS2* has also been linked to early ischemic stroke by GWAS [54, 55]. Neurovascular marker data were not available in most of the datasets included in this study, however we identified a nominally significant association of *ADAMTS2* expression with increased levels of ICAM1, a cell surface glycoprotein that contributes to vascular pathology and the recruitment of inflammatory cells, in FHS brain donor tissue (P=5.57×10^−4^). These findings combined with our observed association of *ADAMTS2* expression with a summarized score for executive function may suggest that increased expression levels of *ADAMTS2* contribute to cognitive impairment through extracellular matrix and vascular dysfunction.

*ADAMTS2* and *SLC6A9* showed a significant trend of increasing expression according to the degree of AD severity (control < AysmAD < SymAD), suggesting the possiblility that their mechanism of action is more closely aligned with the development of the hallmark and perhaps co-morbid pathologies of AD rather than cognitive decline. In fact, expression of *ADAMTS2* and *SLC6A9* was associated with increased levels of several phosphorylated tau residues in both AD cases and controls. Expression of *SLC6A9* was also associated with protein markers of vascular disease, neuroinflammation, and synaptic density in controls and AsymAD cases. Although *SLC6A9* expression was associated with language domain scores, it was not associated with either memory performance or executive function which are stronger indicators of cognitive decline. However, there is other evidence linking *SLC6A9* to psychiatric disturbance [56, 57]. *SLC6A9* encodes GlyT1, a glycine transporter located in the plasma membrane predominantly in glial cells, neurons and astrocytes. GlyT1 modulates extracelluar glycine in the synapse by transporting glycine from the synaptic cleft into cells [56, 57]. Dysregulation of this transproter has been implicated in glycine encephalopathy (a disorder characterized by cognitive impairment) [56], and schizophrenia [58, 59] and depression [60]. Thus, comparison of *SLC6A9* expression in AD cases with and without psychiatric comorbidity may warrant further investigation.

Several other genes that were significantly differentially expressed between AD cases and controls (P<5.0 ×10^−8^) have been implicated in AD or cognitive decline. *SLC38A2*, also known as *SNAT2*, is a sodium-coupled neutral amino acid transporter essential for maintaining amino acid homeostasis. In our study, *SLC38A2* expression was significantly upregulated in the prefrontal cortex of AD cases and strongly associated with language performance. A previous DGE study showed progressive upregulation of *SLC28A2* expression across multiple brain regions in 27 controls, 33 AsymAD cases and 52 SymAD cases from the Medical Research Council London Neurodegenerative Diseases Brain Bank [2], a finding consistent with other studies reporting higher *SLC38A2* expression in AD cases compared to controls [61–63]. The protein encoded by *CHGA* (chromogranin A) has been found at elevated levels in the brains and cerebrospinal fluid (CSF) of AD patients [64, 65] and individuals with MCI, especially those who later progressed to AD [66]. *VAT1* (vesicle amine transport 1) plays a key role in synaptic vesicle function, essential for neurotransmitter release and neuronal communication. Elevated *VAT1* levels have been associated with accelerated cognitive decline, suggesting a complex role in the regulation of neurodegenerative processes [67, 68]. Although the biological relevance of *EMP3* to AD is unclear, similar to our finding its expression was previously shown to be increased in AD compared to control brains [69].

The significant biological pathways identified by gene enrichment analysis involve E2F targets, cholesterol homeostasis, and oxidative phosphorylation, all of which have been linked to AD and neurodegeneration. E2F is a group of genes that encodes a family of transcription factors that regulate processes such as DNA repair, cell cycle control, and apoptosis [70], and may act as key regulatory mediators connecting energy metabolism, autophagy and cellular stress to AD [71]. Abnormalities in lipid metabolism have been demonstrated to promote AD pathogenesis by stimulating Aβ deposition and formation of tau protein tangles [72]. Studies of mitochondrial dynamics in AD brain and mouse models have shown that mitochondrial dysfunction and resultant impaired oxidative phosphorylation affects neurons that are particularly vulnerable to oxidative stress [73].

This study has several limitations. First, some of the autopsy cohorts are relatively small and thus underpowered to detect differential expression between diagnostic subgroups. However, this concern is mitigated by our focus on results emerging from analyses of the total sample combined with the requirement of nominally significant p-values in at least two datasets. Second, associations of gene expression with cognitive domain scores and neuropathological traits were not tested across all four datasets due to the availability of trait data. However, the results were confirmed in at least two datasets. Additionally, our analysis included only participants of European ancestry, which limits the generalizability of our findings. On the other hand, *ADAMTS2* was the top-ranked DEG in both this study and another study conducted in brain tissue from African American AD cases and controls [42]. Finally, the interpretation of our findings is predicated on a construct of cognitive resilience based on the absence of cognitive impairment among persons with a pathologically confirmed diagnosis of AD and this definition may vary according to which clinical and neuropathological variables are considered. Future work should consider the role of alpha-synuclein and TDP-43 inclusions as well as other co-morbid pathologies in gene expression related to cognitive resilience. Cognitive resilience has also been framed according to processes that occur throughout aging which can be measured such as fluid and imaging biomarkers and life experiences including education and occupational attainment [74]. These challenges underscore the need for further research to establish a consensus definition and to elucidate the complex mechanisms underlying cognitive resilience.

## Conclusions

This study identified multiple DEGs that may be involved in cognitive resilience. Our study also illustrated that expression differences of some top-ranked genes are closely associated with performance in multiple cognitive domains and Tau and vascular pathologies. Our findings may provide insight about therapeutic targets for promoting cognitive resilience.

## Ethics approval and consent to participate

The study protocol, design, and performance of the current study were approved by the Boston University Institutional Review Board.

## Consent for publication

Not applicable

## Appendix (Supplementary information)

Supplementary_Table

Supplementary_Figure

## Availability data and materials

**RNA-Seq Data:**

ROSMAP: https://www.synapse.org/Synapse:syn3388564

MSBB: https://www.synapse.org/Synapse:syn3157743

RNA-sequencing data from FHS and BUADRC studies are available upon request.

## Data Availability

RNA-sequencing data from FHS and BUADRC studies are available upon request.

https://www.synapse.org/Synapse:syn3388564

https://www.synapse.org/Synapse:syn3157743

## References

1. Gauthier S, Rosa-Neto P, Morais J, Webster C: World Alzheimer Report 2021: Journey through the diagnosis of dementia. Alzheimer’s Disease International 2021.

2. Patel H, Hodges AK, Curtis C, Lee SH, Troakes C, Dobson RJB, Newhouse SJ: Transcriptomic analysis of probable asymptomatic and symptomatic alzheimer brains. Brain Behav Immun 2019, 80:644–656.

3. Montine TJ, Phelps CH, Beach TG, Bigio EH, Cairns NJ, Dickson DW, Duyckaerts C, Frosch MP, Masliah E, Mirra SS, et al: National Institute on Aging-Alzheimer’s Association guidelines for the neuropathologic assessment of Alzheimer’s disease: a practical approach. Acta Neuropathol 2012, 123:1–11.

4. Ganz AB, Beker N, Hulsman M, Sikkes S, Netherlands Brain B, Scheltens P, Smit AB, Rozemuller AJM, Hoozemans JJM, Holstege H: Neuropathology and cognitive performance in self-reported cognitively healthy centenarians. Acta Neuropathol Commun 2018, 6:64.

5. DeTure MA, Dickson DW: The neuropathological diagnosis of Alzheimer’s disease. Mol Neurodegener 2019, 14:32.

6. Lista S, Zetterberg H, O’Bryant SE, Blennow K, Hampel H: Evolving Relevance of Neuroproteomics in Alzheimer’s Disease. Methods Mol Biol 2017, 1598:101–115.

7. Castrillo JI, Lista S, Hampel H, Ritchie CW: Systems Biology Methods for Alzheimer’s Disease Research Toward Molecular Signatures, Subtypes, and Stages and Precision Medicine: Application in Cohort Studies and Trials. Methods Mol Biol 2018, 1750:31–66.

8. De Strooper B, Karran E: The Cellular Phase of Alzheimer’s Disease. Cell 2016, 164:603–615.

9. Higginbotham L, Ping L, Dammer EB, Duong DM, Zhou M, Gearing M, Hurst C, Glass JD, Factor SA, Johnson ECB, et al: Integrated proteomics reveals brain-based cerebrospinal fluid biomarkers in asymptomatic and symptomatic Alzheimer’s disease. Sci Adv 2020, 6.

10. Caselli RJ, Reiman EM: Characterizing the preclinical stages of Alzheimer’s disease and the prospect of presymptomatic intervention. J Alzheimers Dis 2013, 33 Suppl 1:S405–416.

11. Sperling RA, Aisen PS, Beckett LA, Bennett DA, Craft S, Fagan AM, Iwatsubo T, Jack CR, Jr., Kaye J, Montine TJ, et al: Toward defining the preclinical stages of Alzheimer’s disease: recommendations from the National Institute on Aging-Alzheimer’s Association workgroups on diagnostic guidelines for Alzheimer’s disease. Alzheimers Dement 2011, 7:280–292.

12. Driscoll I, Troncoso J: Asymptomatic Alzheimer’s disease: a prodrome or a state of resilience? Curr Alzheimer Res 2011, 8:330–335.

13. Tsao CW, Vasan RS: Cohort Profile: The Framingham Heart Study (FHS): overview of milestones in cardiovascular epidemiology. Int J Epidemiol 2015, 44:1800–1813.

14. Ashendorf L, Alosco ML, Bing-Canar H, Chapman KR, Martin B, Chaisson CE, Dixon D, Steinberg EG, Tripodis Y, Kowall NW, Stern RA: Clinical Utility of Select Neuropsychological Assessment Battery Tests in Predicting Functional Abilities in Dementia. Arch Clin Neuropsychol 2018, 33:530–540.

15. Galetta KM, Chapman KR, Essis MD, Alosco ML, Gillard D, Steinberg E, Dixon D, Martin B, Chaisson CE, Kowall NW, et al: Screening Utility of the King-Devick Test in Mild Cognitive Impairment and Alzheimer Disease Dementia. Alzheimer Dis Assoc Disord 2017, 31:152–158.

16. De Jager PL, Ma Y, McCabe C, Xu J, Vardarajan BN, Felsky D, Klein HU, White CC, Peters MA, Lodgson B, et al: A multi-omic atlas of the human frontal cortex for aging and Alzheimer’s disease research. Sci Data 2018, 5:180142.

17. Wang M, Beckmann ND, Roussos P, Wang E, Zhou X, Wang Q, Ming C, Neff R, Ma W, Fullard JF, et al: The Mount Sinai cohort of large-scale genomic, transcriptomic and proteomic data in Alzheimer’s disease. Sci Data 2018, 5:180185.

18. Vonsattel JP, Del Amaya MP, Keller CE: Twenty-first century brain banking. Processing brains for research: the Columbia University methods. Acta Neuropathol 2008, 115:509–532.

19. Mez J, Solomon TM, Daneshvar DH, Murphy L, Kiernan PT, Montenigro PH, Kriegel J, Abdolmohammadi B, Fry B, Babcock KJ, et al: Assessing clinicopathological correlation in chronic traumatic encephalopathy: rationale and methods for the UNITE study. Alzheimers Res Ther 2015, 7:62.

20. Friedberg JS, Aytan N, Cherry JD, Xia W, Standring OJ, Alvarez VE, Nicks R, Svirsky S, Meng G, Jun G, et al: Associations between brain inflammatory profiles and human neuropathology are altered based on apolipoprotein E ε4 genotype. Sci Rep 2020, 10:2924.

21. Braak H, Braak E: Neuropathological stageing of Alzheimer-related changes. Acta Neuropathol 1991, 82:239–259.

22. Mirra SS, Heyman A, McKeel D, Sumi SM, Crain BJ, Brownlee LM, Vogel FS, Hughes JP, van Belle G, Berg L: The Consortium to Establish a Registry for Alzheimer’s Disease (CERAD). Part II. Standardization of the neuropathologic assessment of Alzheimer’s disease. Neurology 1991, 41:479–486.

23. Montine TJ, Phelps CH, Beach TG, Bigio EH, Cairns NJ, Dickson DW, Duyckaerts C, Frosch MP, Masliah E, Mirra SS, et al: National Institute on Aging-Alzheimer’s Association guidelines for the neuropathologic assessment of Alzheimer’s disease: a practical approach. Acta Neuropathol 2012, 123:1–11.

24. Panitch R, Hu J, Chung J, Zhu C, Meng G, Xia W, Bennett DA, Lunetta KL, Ikezu T, Au R, et al: Integrative brain transcriptome analysis links complement component 4 and HSPA2 to the APOE ε2 protective effect in Alzheimer disease. Mol Psychiatry 2021.

25. Kang M, Ang TFA, Devine SA, Sherva R, Mukherjee S, Trittschuh EH, Gibbons LE, Scollard P, Lee M, Choi SE, et al: A genome-wide search for pleiotropy in more than 100,000 harmonized longitudinal cognitive domain scores. Mol Neurodegener 2023, 18:40.

26. Mukherjee S, Mez J, Trittschuh EH, Saykin AJ, Gibbons LE, Fardo DW, Wessels M, Bauman J, Moore M, Choi SE, et al: Genetic data and cognitively defined late-onset Alzheimer’s disease subgroups. Mol Psychiatry 2020, 25:2942–2951.

27. Scollard P, Choi SE, Lee ML, Mukherjee S, Trittschuh EH, Sanders RE, Gibbons LE, Joshi P, Devine S, Au R, et al: Ceiling effects and differential measurement precision across calibrated cognitive scores in the Framingham Study. Neuropsychology 2023, 37:383–397.

28. Mukherjee S, Choi SE, Lee ML, Scollard P, Trittschuh EH, Mez J, Saykin AJ, Gibbons LE, Sanders RE, Zaman AF, et al: Cognitive domain harmonization and cocalibration in studies of older adults. Neuropsychology 2023, 37:409–423.

29. Muthén LK, Muthén BO: Mplus: Statistical analysis with latent variables; user’s guide;[version 7]. Muthén et Muthén; 2012.

30. Bachstetter AD, Van Eldik LJ, Schmitt FA, Neltner JH, Ighodaro ET, Webster SJ, Patel E, Abner EL, Kryscio RJ, Nelson PT: Disease-related microglia heterogeneity in the hippocampus of Alzheimer’s disease, dementia with Lewy bodies, and hippocampal sclerosis of aging. Acta Neuropathol Commun 2015, 3:32.

31. Cherry JD, Tripodis Y, Alvarez VE, Huber B, Kiernan PT, Daneshvar DH, Mez J, Montenigro PH, Solomon TM, Alosco ML, et al: Microglial neuroinflammation contributes to tau accumulation in chronic traumatic encephalopathy. Acta Neuropathol Commun 2016, 4:112.

32. Panitch R, Hu J, Chung J, Zhu C, Meng G, Xia W, Bennett DA, Lunetta KL, Ikezu T, Au R, et al: Integrative brain transcriptome analysis links complement component 4 and HSPA2 to the APOE epsilon2 protective effect in Alzheimer disease. Mol Psychiatry 2021, 26:6054–6064.

33. Stathas S, Alvarez VE, Xia W, Nicks R, Meng G, Daley S, Pothast M, Shah A, Kelley H, Esnault C, et al: Tau phosphorylation sites serine202 and serine396 are differently altered in chronic traumatic encephalopathy and Alzheimer’s disease. Alzheimers Dement 2021.

34. Risso D, Ngai J, Speed TP, Dudoit S: Normalization of RNA-seq data using factor analysis of control genes or samples. Nat Biotechnol 2014, 32:896–902.

35. Love MI, Huber W, Anders S: Moderated estimation of fold change and dispersion for RNA-seq data with DESeq2. Genome Biol 2014, 15:550.

36. Willer CJ, Li Y, Abecasis GR: METAL: fast and efficient meta-analysis of genomewide association scans. Bioinformatics 2010, 26:2190–2191.

37. Chen EY, Tan CM, Kou Y, Duan Q, Wang Z, Meirelles GV, Clark NR, Ma’ayan A: Enrichr: interactive and collaborative HTML5 gene list enrichment analysis tool. BMC Bioinformatics 2013, 14:128.

38. Subramanian A, Tamayo P, Mootha VK, Mukherjee S, Ebert BL, Gillette MA, Paulovich A, Pomeroy SL, Golub TR, Lander ES, Mesirov JP: Gene set enrichment analysis: a knowledge-based approach for interpreting genome-wide expression profiles. Proc Natl Acad Sci U S A 2005, 102:15545–15550.

39. Liberzon A, Birger C, Thorvaldsdottir H, Ghandi M, Mesirov JP, Tamayo P: The Molecular Signatures Database (MSigDB) hallmark gene set collection. Cell Syst 2015, 1:417–425.

40. Kanehisa M, Goto S: KEGG: kyoto encyclopedia of genes and genomes. Nucleic Acids Res 2000, 28:27–30.

41. Jun G, Guo H, Klein BE, Klein R, Wang JJ, Mitchell P, Miao H, Lee KE, Joshi T, Buck M, et al: EPHA2 is associated with age-related cortical cataract in mice and humans. PLoS Genet 2009, 5:e1000584.

42. Logue M, Labadorf A, O’Niell NK, Dickson DW, Dugger BN, Flanagan ME, Frosch MP, Gearing M, Jin L-W, Kofler J, et al: Transcriptome-wide association study of Alzheimer disease reveals many differentially expressed genes and multiple biological pathways in brain tissue from African American donors. medRxiv 2024:2024.2010.2029.24316311.

43. McCorkindale AN, Patrick E, Duce JA, Guennewig B, Sutherland GT: The Key Factors Predicting Dementia in Individuals With Alzheimer’s Disease-Type Pathology. Front Aging Neurosci 2022, 14:831967.

44. Bellenguez C, Küçükali F, Jansen IE, Kleineidam L, Moreno-Grau S, Amin N, Naj AC, Campos-Martin R, Grenier-Boley B, Andrade V, et al: New insights into the genetic etiology of Alzheimer’s disease and related dementias. Nat Genet 2022, 54:412–436.

45. Tang BL, Hong W: ADAMTS: a novel family of proteases with an ADAM protease domain and thrombospondin 1 repeats. FEBS Lett 1999, 445:223–225.

46. Colige A, Nuytinck L, Hausser I, van Essen AJ, Thiry M, Herens C, Ades LC, Malfait F, Paepe AD, Franck P, et al: Novel types of mutation responsible for the dermatosparactic type of Ehlers-Danlos syndrome (Type VIIC) and common polymorphisms in the ADAMTS2 gene. J Invest Dermatol 2004, 123:656–663.

47. Bekhouche M, Colige A: The procollagen N-proteinases ADAMTS2, 3 and 14 in pathophysiology. Matrix Biol 2015, 44-46:46–53.

48. Soles A, Selimovic A, Sbrocco K, Ghannoum F, Hamel K, Moncada EL, Gilliat S, Cvetanovic M: Extracellular Matrix Regulation in Physiology and in Brain Disease. Int J Mol Sci 2023, 24.

49. Anwar MM, Ozkan E, Gursoy-Ozdemir Y: The role of extracellular matrix alterations in mediating astrocyte damage and pericyte dysfunction in Alzheimer’s disease: A comprehensive review. Eur J Neurosci 2022, 56:5453–5475.

50. Lussier AL, Weeber EJ, Rebeck GW: Reelin Proteolysis Affects Signaling Related to Normal Synapse Function and Neurodegeneration. Front Cell Neurosci 2016, 10:75.

51. Bock HH, May P: Canonical and Non-canonical Reelin Signaling. Frontiers in Cellular Neuroscience 2016, 10.

52. Bakota L, Brandt R: Tau Biology and Tau-Directed Therapies for Alzheimer’s Disease. Drugs 2016, 76:301–313.

53. Sun Y, Xu S, Jiang M, Liu X, Yang L, Bai Z, Yang Q: Role of the Extracellular Matrix in Alzheimer’s Disease. Front Aging Neurosci 2021, 13:707466.

54. Earley EJ, Kelly S, Fang F, Alencar CS, Rodrigues DOW, Soares Cruz DT, Flanagan JM, Ware RE, Zhang X, Gordeuk V, et al: Genome-wide association study of early ischaemic stroke risk in Brazilian individuals with sickle cell disease implicates ADAMTS2 and CDK18 and uncovers novel loci. Br J Haematol 2023, 201:343–352.

55. Arning A, Hiersche M, Witten A, Kurlemann G, Kurnik K, Manner D, Stoll M, Nowak-Gottl U: A genome-wide association study identifies a gene network of ADAMTS genes in the predisposition to pediatric stroke. Blood 2012, 120:5231–5236.

56. Ayka A, Sehirli AO: The Role of the SLC Transporters Protein in the Neurodegenerative Disorders. Clin Psychopharmacol Neurosci 2020, 18:174–187.

57. Harvey RJ, Yee BK: Glycine transporters as novel therapeutic targets in schizophrenia, alcohol dependence and pain. Nat Rev Drug Discov 2013, 12:866–885.

58. Kristensen AS, Andersen J, Jorgensen TN, Sorensen L, Eriksen J, Loland CJ, Stromgaard K, Gether U: SLC6 neurotransmitter transporters: structure, function, and regulation. Pharmacol Rev 2011, 63:585–640.

59. Broer S, Gether U: The solute carrier 6 family of transporters. Br J Pharmacol 2012, 167:256–278.

60. Bai X, Moraes TF, Reithmeier RAF: Structural biology of solute carrier (SLC) membrane transport proteins. Mol Membr Biol 2017, 34:1–32.

61. Li Y, Shi H, Chen T, Xue J, Wang C, Peng M, Si G: Establishing a competing endogenous RNA (ceRNA)-immunoregulatory network associated with the progression of Alzheimer’s disease. Ann Transl Med 2022, 10:65.

62. Wang XL, Li L: Cell type-specific potential pathogenic genes and functional pathways in Alzheimer’s Disease. BMC Neurol 2021, 21:381.

63. Santiago JA, Bottero V, Potashkin JA: Transcriptomic and Network Analysis Identifies Shared and Unique Pathways across Dementia Spectrum Disorders. Int J Mol Sci 2020, 21.

64. Sathe G, Albert M, Darrow J, Saito A, Troncoso J, Pandey A, Moghekar A: Quantitative proteomic analysis of the frontal cortex in Alzheimer’s disease. J Neurochem 2021, 156:988–1002.

65. Quinn JP, Ethier EC, Novielli A, Malone A, Ramirez CE, Salloum L, Trombetta BA, Kivisakk P, Bremang M, Selzer S, et al: Cerebrospinal Fluid and Brain Proteoforms of the Granin Neuropeptide Family in Alzheimer’s Disease. J Am Soc Mass Spectrom 2023, 34:649–667.

66. Duits FH, Brinkmalm G, Teunissen CE, Brinkmalm A, Scheltens P, Van der Flier WM, Zetterberg H, Blennow K: Synaptic proteins in CSF as potential novel biomarkers for prognosis in prodromal Alzheimer’s disease. Alzheimers Res Ther 2018, 10:5.

67. Zammit AR, Yu L, Petyuk V, Schneider JA, De Jager PL, Klein HU, Bennett DA, Buchman AS: Cortical Proteins and Individual Differences in Cognitive Resilience in Older Adults. Neurology 2022, 98:e1304–e1314.

68. de Vries LE, Huitinga I, Kessels HW, Swaab DF, Verhaagen J: The concept of resilience to Alzheimer’s Disease: current definitions and cellular and molecular mechanisms. Mol Neurodegener 2024, 19:33.

69. Ramos-Campoy O, Llado A, Bosch B, Ferrer M, Perez-Millan A, Vergara M, Molina-Porcel L, Fort-Aznar L, Gonzalo R, Moreno-Izco F, et al: Differential Gene Expression in Sporadic and Genetic Forms of Alzheimer’s Disease and Frontotemporal Dementia in Brain Tissue and Lymphoblastoid Cell Lines. Mol Neurobiol 2022, 59:6411–6428.

70. Greene LA, Biswas SC, Liu DX: Cell cycle molecules and vertebrate neuron death: E2F at the hub. Cell Death Differ 2004, 11:49–60.

71. Casey AE, Liu W, Hein LK, Sargeant TJ, Pederson SM, Makinen VP: Transcriptional targets of senataxin and E2 promoter binding factors are associated with neuro-degenerative pathways during increased autophagic flux. Sci Rep 2022, 12:17665.

72. Tong B, Ba Y, Li Z, Yang C, Su K, Qi H, Zhang D, Liu X, Wu Y, Chen Y, et al: Targeting dysregulated lipid metabolism for the treatment of Alzheimer’s disease and Parkinson’s disease: Current advancements and future prospects. Neurobiol Dis 2024, 196:106505.

73. Zhu X, Perry G, Smith MA, Wang X: Abnormal mitochondrial dynamics in the pathogenesis of Alzheimer’s disease. J Alzheimers Dis 2013, 33 Suppl 1:S253–262.

74. Joshi MS, Galvin JE: Cognitive Resilience in Brain Health and Dementia Research. J Alzheimers Dis 2022, 90:461–473.

