## Supplementary Figure for "Transcriptome Signatures for Cognitive Resilience Among Individuals with Pathologically Confirmed Alzheimer Disease"

**Supplementary Figure 1:** Enrichment analysis seeded with 4,266 genes that were differentially expressed ($P < 0.05$) between SymAD and AsymAD cases in the total sample. Pathways were identified using the **(A)** Molecular Signatures 2020 and **(B)** KEGG Human 2021 databases. Pathway enrichment P-values are indicated according to the color-coded key beside each panel and were FDR-adjusted. The x-axis indicates the number of DEGs in each pathway.

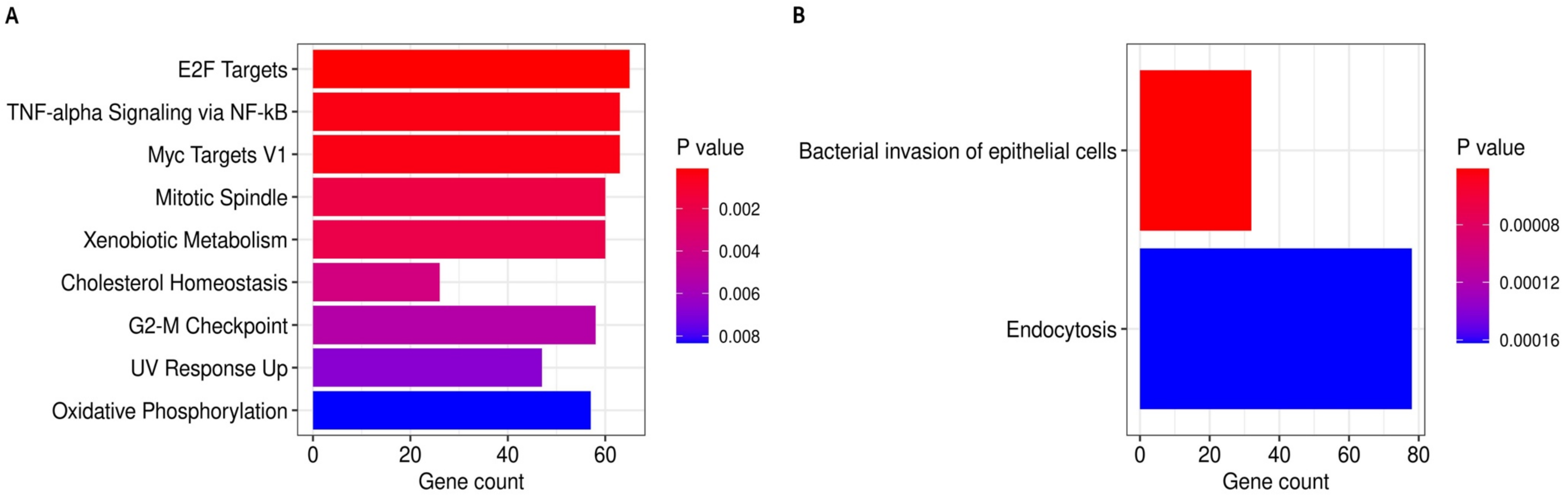
