## Supplementary Tables for "Transcriptome Signatures for Cognitive Resilience Among Individuals with Pathologically Confirmed Alzheimer Disease"

**Supplementary Table 1:** Summary of cognitive scores in SymAD, AsymAD, and Control subjects from the ROSMAP, FHS, and BUADRC datasets.

| Grp | Domain | ROSMAP | | | | | | | | FHS | | | | | | | | BUADRC | | | | | | | |
|---|---|---|---|---|---|---|---|---|---|---|---|---|---|---|---|---|---|---|---|---|---|---|---|---|---|
| | | All (n=567) | | SymAD (n=200) | | AsymAD (n=172) | | Control (n=195) | | All (n=94) | | SymAD (n=30) | | AsymAD (n=9) | | Control (n=55) | | All (n=85) | | SymAD (n=30) | | AsymAD (n=20) | | Control (n=35) | |
| | | Mean | SD | Mean | SD | Mean | SD | Mean | SD | Mean | SD | Mean | SD | Mean | SD | Mean | SD | Mean | SD | Mean | SD | Mean | SD | Mean | SD |
| 1 | **Memory** | -0.490 | 0.995 | -1.574 | 0.640 | -0.074 | 0.541 | 0.227 | 0.583 | 0.031 | 0.775 | -0.901 | 0.613 | 0.320 | 0.533 | 0.407 | 0.459 | -0.473 | 1.316 | -1.523 | 0.842 | -0.555 | 1.258 | 0.444 | 0.973 |
| 2 | **Language** | -0.496 | 0.939 | -1.480 | 0.732 | -0.133 | 0.549 | 0.129 | 0.536 | -0.091 | 0.678 | -0.740 | 0.316 | -0.160 | 0.747 | 0.173 | 0.599 | -0.405 | 1.058 | -1.224 | 0.706 | -0.487 | 1.050 | 0.273 | 0.812 |
| 3 | **Executive Function** | -0.239 | 0.757 | -1.037 | 0.588 | 0.008 | 0.507 | 0.231 | 0.474 | -0.255 | 0.590 | -0.690 | 0.577 | -0.373 | 0.500 | -0.078 | 0.527 | -0.560 | 0.859 | -1.264 | 0.742 | -0.588 | 0.668 | -0.207 | 0.784 |

**Supplementary Table 2:** Summary of neuropathological traits in SymAD, AsymAD, and Control subjects from the FHS and BUADRC datasets.

| Grp | Pathology Group | Traits | FHS | | | | | | | | BUADRC | | | | | | | |
|---|---|---|---|---|---|---|---|---|---|---|---|---|---|---|---|---|---|---|
| | | | All (n=127) | | SymAD (n=42) | | AsymAD (n=12) | | Control (n=73) | | All (n=85) | | SymAD (n=30) | | AsymAD (n=20) | | Control (n=35) | |
| | | | Mean/ Median | SD/ Range | Mean/ Median | SD/ Range | Mean/ Median | SD/ Range | Mean/ Median | SD/ Range | Mean/ Median | SD/ Range | Mean/ Median | SD/ Range | Mean/ Median | SD/ Range | Mean/ Median | SD/ Range |
| 1 | Tau pathology | **Braak Stage (score 0-6)** | 3 | 0~6 | 5 | 3~6 | 4 | 3~6 | 2 | 0~4 | 4 | 0~6 | 6 | 3~6 | 5 | 3~6 | 3 | 0~4 |
| 1 | Tau pathology | **AT8(positive pixels/mm2)** | 18496 | 56319 | 50247 | 88005 | 9639 | 19138 | 653 | 589 | 28521 | 55638 | 55790 | 74930 | 36995 | 51754 | 1329 | 3949 |
| 1 | Tau pathology | **pTau181 (U/g)** | 11842 | 6123 | 13868 | 6827 | 10192 | 4993 | 10948 | 5622 | 62962 | 44746 | 74314 | 43707 | 72104 | 41152 | 48007 | 44473 |
| 1 | Tau pathology | **pTau231 (U/g)** | 623238 | 588276 | 916088 | 863556 | 423253 | 292265 | 483804 | 291200 | 1326834 | 1903678 | 2380647 | 2747745 | 1169680 | 1102169 | 513368 | 398950 |
| 1 | Tau pathology | **pTau202 (U/g)** | 2109070 | 4684939 | 4071206 | 7174011 | 2063201 | 3154167 | 967682 | 1943305 | 22905392 | 51094133 | 50155669 | 72070867 | 21682755 | 40854672 | 990308 | 2518545 |
| 1 | Tau pathology | **pTau396 (U/g)** | 14786 | 28709 | 26114 | 40218 | 14424 | 24473 | 8213 | 17438 | 90754 | 221858 | 191711 | 324550 | 90138 | 174610 | 7456 | 23206 |
| 2 | Beta-amyloid pathology | **CERAD (score 0-3)** | 1 | 0~3 | 3 | 1~3 | 2.5 | 1~3 | 0 | 0~3 | 2 | 0~3 | 3 | 1~3 | 2 | 1~3 | 0 | 0~3 |
| 2 | Beta-amyloid pathology | **Ab40 (pg/mg)** | 328 | 794 | 448 | 839 | 861 | 1256 | 171 | 616 | 835 | 1939 | 1633 | 2568 | 995 | 2100 | 61 | 90 |
| 2 | Beta-amyloid pathology | **Ab42 (pg/mg)** | 1043 | 911 | 1333 | 846 | 1605 | 1259 | 784 | 795 | 1122 | 1153 | 1488 | 909 | 1414 | 1497 | 641 | 961 |
| 3 | Synaptic density | **PSD-95 (U/g)** | 48918 | 27735 | 44665 | 25269 | 44927 | 27978 | 52063 | 28997 | 53885 | 27208 | 62519 | 25364 | 49256 | 31792 | 49131 | 24774 |
| 3 | Synaptic density | **Asyn (alpha-synuclein) (ug/ml)** | 15 | 7 | 14 | 6 | 16 | 3 | 15 | 8 | 12 | 5 | 13 | 5 | 11 | 4 | 11 | 3 |
| 4 | Neuroinflammatory | **Iba1 (cells/$mm^2$)** | 179 | 46 | 197 | 47 | 187 | 39 | 168 | 43 | 171 | 46 | 157 | 52 | 179 | 34 | 178 | 46 |

**Supplementary Table 3:** Meta-analysis results of differential expression between SymAD and AsymAD across four datasets. DEGs at the TWS level (P<10e-6) in meta-analysis and those that were nominally significant (P<0.05) in at least two datasets were listed. The genes in bold indicate the meta P-value < 5e-8.

| | Meta-analysis | | | | ROSMP | | | MSBB | | | FHS | | | BUADRC | | |
|---|---|---|---|---|---|---|---|---|---|---|---|---|---|---|---|---|
| Gene | Log2FC | LfcSE | P.value | Direction | Log2FC | LfcSE | P.value | Log2FC | LfcSE | P.value | Log2FC | LfcSE | P.value | Log2FC | LfcSE | P.value |
| ***ADAMTS2*** | 0.458 | 0.060 | 2.94E-14 | ++++ | 0.421 | 0.070 | 2.16E-09 | 0.628 | 0.313 | 4.49E-02 | 0.542 | 0.194 | 5.12E-03 | 0.556 | 0.166 | 8.20E-04 |
| ***S100A4*** | 0.605 | 0.092 | 3.98E-11 | ++++ | 0.530 | 0.106 | 5.44E-07 | 1.032 | 0.493 | 3.64E-02 | 0.840 | 0.265 | 1.51E-03 | 0.748 | 0.297 | 1.17E-02 |
| ***NRIP2*** | 0.320 | 0.050 | 9.52E-11 | ++++ | 0.295 | 0.055 | 7.78E-08 | 0.365 | 0.205 | 7.40E-02 | 0.527 | 0.221 | 1.73E-02 | 0.419 | 0.176 | 1.75E-02 |
| ***SCGN*** | 0.548 | 0.087 | 2.55E-10 | ++++ | 0.480 | 0.097 | 6.90E-07 | 0.895 | 0.405 | 2.72E-02 | 0.897 | 0.341 | 8.48E-03 | 0.732 | 0.294 | 1.26E-02 |
| ***SLC38A2*** | 0.308 | 0.049 | 4.71E-10 | ++++ | 0.289 | 0.056 | 2.66E-07 | 0.434 | 0.262 | 9.70E-02 | 0.338 | 0.216 | 1.18E-01 | 0.366 | 0.132 | 5.61E-03 |
| ***ALDH1A1*** | -0.333 | 0.054 | 4.90E-10 | ---- | -0.314 | 0.060 | 1.67E-07 | -0.192 | 0.252 | 4.46E-01 | -0.537 | 0.194 | 5.68E-03 | -0.402 | 0.184 | 2.91E-02 |
| ***CHGA*** | 0.219 | 0.036 | 1.35E-09 | ++++ | 0.200 | 0.039 | 1.99E-07 | 0.143 | 0.220 | 5.16E-01 | 0.447 | 0.185 | 1.58E-02 | 0.372 | 0.147 | 1.13E-02 |
| ***CEP83*** | -0.157 | 0.027 | 5.23E-09 | ---- | -0.136 | 0.030 | 4.53E-06 | -0.137 | 0.117 | 2.41E-01 | -0.312 | 0.105 | 2.94E-03 | -0.279 | 0.106 | 8.75E-03 |
| ***VAT1*** | 0.139 | 0.024 | 5.85E-09 | ++++ | 0.130 | 0.026 | 8.47E-07 | 0.395 | 0.155 | 1.09E-02 | 0.078 | 0.090 | 3.83E-01 | 0.208 | 0.082 | 1.14E-02 |
| ***PAFAH1B3*** | 0.144 | 0.025 | 1.20E-08 | ++++ | 0.147 | 0.028 | 1.36E-07 | 0.029 | 0.141 | 8.35E-01 | 0.117 | 0.101 | 2.47E-01 | 0.172 | 0.084 | 4.07E-02 |
| ***POLD1*** | 0.158 | 0.028 | 1.57E-08 | ++++ | 0.153 | 0.031 | 6.54E-07 | 0.241 | 0.196 | 2.19E-01 | 0.141 | 0.094 | 1.34E-01 | 0.222 | 0.110 | 4.47E-02 |
| ***IVD*** | -0.106 | 0.019 | 2.13E-08 | ---- | -0.100 | 0.020 | 8.58E-07 | -0.088 | 0.094 | 3.49E-01 | -0.258 | 0.110 | 1.85E-02 | -0.127 | 0.075 | 9.14E-02 |
| ***ARG2*** | 0.275 | 0.050 | 2.82E-08 | ++++ | 0.285 | 0.057 | 4.88E-07 | 0.206 | 0.174 | 2.37E-01 | 0.384 | 0.191 | 4.45E-02 | 0.169 | 0.170 | 3.21E-01 |
| ***SLC6A9*** | 0.239 | 0.043 | 2.93E-08 | ++++ | 0.220 | 0.048 | 4.85E-06 | 0.564 | 0.273 | 3.87E-02 | 0.156 | 0.161 | 3.35E-01 | 0.364 | 0.134 | 6.66E-03 |
| *TMCC2* | 0.149 | 0.027 | 5.06E-08 | ++++ | 0.153 | 0.031 | 5.33E-07 | 0.090 | 0.144 | 5.31E-01 | 0.298 | 0.139 | 3.19E-02 | 0.093 | 0.078 | 2.36E-01 |
| *CLIP2* | 0.135 | 0.025 | 6.01E-08 | ++++ | 0.131 | 0.028 | 3.36E-06 | 0.127 | 0.091 | 1.61E-01 | 0.159 | 0.110 | 1.49E-01 | 0.165 | 0.084 | 4.80E-02 |
| *ELP2* | -0.067 | 0.013 | 7.27E-08 | --+- | -0.072 | 0.014 | 1.33E-07 | -0.149 | 0.063 | 1.84E-02 | 0.103 | 0.058 | 7.64E-02 | -0.080 | 0.047 | 8.84E-02 |
| *ARSG* | -0.146 | 0.027 | 7.80E-08 | ---- | -0.132 | 0.030 | 9.90E-06 | -0.124 | 0.126 | 3.27E-01 | -0.239 | 0.116 | 4.01E-02 | -0.241 | 0.099 | 1.45E-02 |
| *QDPR* | 0.224 | 0.042 | 1.02E-07 | ++++ | 0.182 | 0.046 | 7.15E-05 | 0.492 | 0.217 | 2.35E-02 | 0.464 | 0.207 | 2.52E-02 | 0.427 | 0.152 | 4.83E-03 |
| *NSMCE1* | -0.103 | 0.019 | 1.03E-07 | ---- | -0.104 | 0.021 | 1.16E-06 | -0.092 | 0.109 | 3.98E-01 | -0.154 | 0.071 | 2.95E-02 | -0.045 | 0.072 | 5.27E-01 |
| *IL2RG* | 0.394 | 0.074 | 1.04E-07 | ++++ | 0.338 | 0.082 | 4.07E-05 | 0.653 | 0.649 | 3.14E-01 | 0.368 | 0.248 | 1.39E-01 | 0.857 | 0.242 | 3.98E-04 |
| *SCG3* | -0.166 | 0.032 | 1.34E-07 | ---- | -0.153 | 0.034 | 8.00E-06 | -0.098 | 0.135 | 4.71E-01 | -0.531 | 0.158 | 7.76E-04 | -0.172 | 0.129 | 1.81E-01 |
| *CXCR4* | 0.423 | 0.080 | 1.42E-07 | ++-+ | 0.458 | 0.093 | 7.99E-07 | 0.668 | 0.424 | 1.16E-01 | -0.192 | 0.258 | 4.57E-01 | 0.631 | 0.235 | 7.13E-03 |
| *RABGGTA* | 0.116 | 0.022 | 1.44E-07 | ++++ | 0.116 | 0.024 | 2.15E-06 | 0.115 | 0.133 | 3.90E-01 | 0.224 | 0.094 | 1.79E-02 | 0.062 | 0.070 | 3.75E-01 |
| *RAB11FIP5* | 0.119 | 0.023 | 1.77E-07 | +-++ | 0.130 | 0.026 | 3.51E-07 | -0.023 | 0.100 | 8.14E-01 | 0.197 | 0.097 | 4.36E-02 | 0.061 | 0.073 | 4.02E-01 |
| *HSD11B2* | 0.409 | 0.079 | 1.98E-07 | +-++ | 0.348 | 0.089 | 1.00E-04 | -0.211 | 0.400 | 5.99E-01 | 0.819 | 0.293 | 5.16E-03 | 0.770 | 0.231 | 8.76E-04 |
| *MON1B* | -0.071 | 0.014 | 2.05E-07 | -+-- | -0.074 | 0.015 | 1.00E-06 | 0.059 | 0.078 | 4.50E-01 | -0.062 | 0.062 | 3.15E-01 | -0.082 | 0.039 | 3.64E-02 |
| *PEPD* | 0.098 | 0.019 | 2.16E-07 | ++-+ | 0.100 | 0.020 | 9.70E-07 | 0.232 | 0.112 | 3.72E-02 | -0.021 | 0.086 | 8.07E-01 | 0.095 | 0.069 | 1.69E-01 |
| *SAP30L* | 0.129 | 0.025 | 2.25E-07 | ++++ | 0.120 | 0.028 | 1.84E-05 | 0.362 | 0.163 | 2.68E-02 | 0.087 | 0.100 | 3.87E-01 | 0.162 | 0.070 | 2.09E-02 |
| *ZBBX* | -0.270 | 0.053 | 2.73E-07 | ---- | -0.260 | 0.058 | 8.17E-06 | -0.352 | 0.235 | 1.34E-01 | -0.182 | 0.229 | 4.27E-01 | -0.376 | 0.181 | 3.82E-02 |
| *USP31* | 0.149 | 0.029 | 2.85E-07 | ++++ | 0.134 | 0.033 | 3.96E-05 | 0.091 | 0.143 | 5.25E-01 | 0.301 | 0.178 | 8.97E-02 | 0.222 | 0.078 | 4.24E-03 |
| *PRELP* | 0.225 | 0.044 | 2.90E-07 | ++++ | 0.243 | 0.051 | 1.78E-06 | 0.556 | 0.262 | 3.36E-02 | 0.180 | 0.160 | 2.61E-01 | 0.099 | 0.111 | 3.69E-01 |
| *FHIP1B* | -0.121 | 0.024 | 2.95E-07 | ---- | -0.110 | 0.027 | 3.58E-05 | -0.121 | 0.090 | 1.78E-01 | -0.247 | 0.093 | 8.08E-03 | -0.130 | 0.084 | 1.22E-01 |
| *MARCHF8* | 0.113 | 0.022 | 3.17E-07 | ++++ | 0.103 | 0.025 | 3.92E-05 | 0.082 | 0.097 | 3.99E-01 | 0.095 | 0.104 | 3.60E-01 | 0.188 | 0.061 | 2.08E-03 |
| *MED29* | -0.127 | 0.025 | 3.29E-07 | ---- | -0.117 | 0.027 | 1.99E-05 | -0.036 | 0.140 | 7.98E-01 | -0.395 | 0.125 | 1.60E-03 | -0.132 | 0.076 | 8.35E-02 |
| *SLC43A3* | 0.204 | 0.040 | 3.88E-07 | ++-+ | 0.219 | 0.045 | 1.22E-06 | 0.628 | 0.306 | 4.03E-02 | -0.088 | 0.138 | 5.26E-01 | 0.258 | 0.125 | 3.88E-02 |
| *ANP32E* | -0.130 | 0.026 | 4.26E-07 | ---- | -0.118 | 0.029 | 5.08E-05 | 0.000 | 0.182 | 9.98E-01 | -0.307 | 0.089 | 6.09E-04 | -0.106 | 0.074 | 1.50E-01 |
| *LRATD2* | 0.167 | 0.033 | 4.93E-07 | ++++ | 0.127 | 0.037 | 6.65E-04 | 0.327 | 0.209 | 1.17E-01 | 0.340 | 0.132 | 1.00E-02 | 0.299 | 0.094 | 1.57E-03 |
| *PRR13* | -0.094 | 0.019 | 5.00E-07 | ---- | -0.099 | 0.021 | 2.98E-06 | -0.030 | 0.133 | 8.23E-01 | -0.222 | 0.073 | 2.49E-03 | -0.011 | 0.053 | 8.31E-01 |
| *PPM1D* | 0.131 | 0.026 | 5.55E-07 | ++++ | 0.123 | 0.029 | 2.22E-05 | 0.128 | 0.126 | 3.12E-01 | 0.104 | 0.124 | 4.01E-01 | 0.195 | 0.079 | 1.33E-02 |
| *MTMR10* | 0.118 | 0.024 | 5.67E-07 | ++-+ | 0.114 | 0.026 | 1.55E-05 | 0.314 | 0.147 | 3.25E-02 | -0.048 | 0.101 | 6.33E-01 | 0.173 | 0.067 | 9.51E-03 |
| *WBP2NL* | -0.256 | 0.051 | 5.67E-07 | ---- | -0.225 | 0.058 | 1.11E-04 | -0.373 | 0.230 | 1.05E-01 | -0.172 | 0.181 | 3.42E-01 | -0.493 | 0.160 | 2.04E-03 |
| *STAG1* | 0.097 | 0.019 | 5.70E-07 | ++-+ | 0.092 | 0.021 | 1.23E-05 | 0.146 | 0.090 | 1.05E-01 | -0.007 | 0.104 | 9.47E-01 | 0.191 | 0.077 | 1.36E-02 |
| *GEM* | 0.379 | 0.076 | 5.92E-07 | ++-+ | 0.444 | 0.090 | 7.35E-07 | 1.015 | 0.359 | 4.64E-03 | -0.035 | 0.266 | 8.96E-01 | 0.117 | 0.191 | 5.42E-01 |
| *MYOT* | 0.265 | 0.053 | 6.06E-07 | ++++ | 0.212 | 0.062 | 5.88E-04 | 0.238 | 0.174 | 1.73E-01 | 0.894 | 0.250 | 3.51E-04 | 0.371 | 0.151 | 1.43E-02 |
| *CHN2* | 0.187 | 0.038 | 6.53E-07 | ++++ | 0.165 | 0.040 | 4.14E-05 | 0.336 | 0.182 | 6.50E-02 | 0.418 | 0.206 | 4.29E-02 | 0.264 | 0.155 | 8.85E-02 |
| *SMPX* | 0.252 | 0.051 | 6.99E-07 | ++++ | 0.204 | 0.054 | 1.63E-04 | 0.345 | 0.263 | 1.90E-01 | 0.669 | 0.259 | 9.69E-03 | 0.777 | 0.244 | 1.45E-03 |
| *PRDM8* | 0.132 | 0.027 | 7.27E-07 | ++++ | 0.100 | 0.030 | 8.56E-04 | 0.095 | 0.163 | 5.60E-01 | 0.258 | 0.123 | 3.52E-02 | 0.266 | 0.070 | 1.33E-04 |
| *EMP3* | 0.212 | 0.043 | 8.06E-07 | ++++ | 0.215 | 0.047 | 5.21E-06 | 0.777 | 0.289 | 7.10E-03 | 0.046 | 0.146 | 7.51E-01 | 0.204 | 0.174 | 2.42E-01 |
| *DYNLRB1* | 0.062 | 0.013 | 8.63E-07 | ++++ | 0.061 | 0.013 | 5.87E-06 | 0.006 | 0.111 | 9.58E-01 | 0.136 | 0.051 | 7.53E-03 | 0.005 | 0.055 | 9.31E-01 |
| *CNKSR3* | 0.172 | 0.035 | 9.63E-07 | ++++ | 0.149 | 0.038 | 8.96E-05 | 0.672 | 0.201 | 8.14E-04 | 0.371 | 0.139 | 7.46E-03 | 0.021 | 0.142 | 8.80E-01 |
| *TNFRSF18* | -0.330 | 0.067 | 9.84E-07 | --+- | -0.359 | 0.076 | 2.42E-06 | -0.138 | 0.469 | 7.68E-01 | 0.158 | 0.329 | 6.30E-01 | -0.342 | 0.173 | 4.75E-02 |

**Supplementary Table 4:** Comparison of DEGs from SymAD and AsymAD differential expression model with BRAAK, CERAD, and APOE4 adjusted models.

| | Base (Sym&AsymAD) | | | Base+BRAAK | | | Base+CERAD | | | Base+BRAAK+CERAD | | | Base+APOE4 | | |
|---|---|---|---|---|---|---|---|---|---|---|---|---|---|---|---|
| **Gene** | **Log2FC** | **LfcSE** | **P.value** | **Log2FC** | **LfcSE** | **P.value** | **Log2FC** | **LfcSE** | **P.value** | **Log2FC** | **LfcSE** | **P.value** | **Log2FC** | **LfcSE** | **P.value** |
| *ADAMTS2* | 0.458 | 0.060 | 2.94E-14 | 0.433 | 0.063 | 5.52E-12 | 0.438 | 0.064 | 8.29E-12 | 0.408 | 0.065 | 3.03E-10 | 0.464 | 0.063 | 2.40E-13 |
| *S100A4* | 0.605 | 0.092 | 3.98E-11 | 0.460 | 0.093 | 7.04E-07 | 0.508 | 0.094 | 6.29E-08 | 0.433 | 0.095 | 4.74E-06 | 0.549 | 0.093 | 3.32E-09 |
| *NRIP2* | 0.320 | 0.050 | 9.52E-11 | 0.267 | 0.052 | 3.43E-07 | 0.263 | 0.053 | 6.39E-07 | 0.233 | 0.053 | 1.26E-05 | 0.304 | 0.053 | 8.05E-09 |
| *SCGN* | 0.548 | 0.087 | 2.55E-10 | 0.495 | 0.088 | 2.12E-08 | 0.475 | 0.088 | 7.83E-08 | 0.452 | 0.090 | 5.38E-07 | 0.494 | 0.088 | 1.73E-08 |
| *SLC38A2* | 0.308 | 0.049 | 4.71E-10 | 0.279 | 0.051 | 5.55E-08 | 0.285 | 0.052 | 4.65E-08 | 0.257 | 0.053 | 1.19E-06 | 0.327 | 0.052 | 2.47E-10 |
| *ALDH1A1* | -0.333 | 0.054 | 4.90E-10 | -0.312 | 0.056 | 2.53E-08 | -0.314 | 0.056 | 2.57E-08 | -0.302 | 0.058 | 1.51E-07 | -0.318 | 0.056 | 1.03E-08 |
| *CHGA* | 0.219 | 0.036 | 1.35E-09 | 0.192 | 0.039 | 8.95E-07 | 0.197 | 0.039 | 4.39E-07 | 0.189 | 0.040 | 2.19E-06 | 0.198 | 0.039 | 2.78E-07 |
| *CEP83* | -0.157 | 0.027 | 5.23E-09 | -0.145 | 0.027 | 1.20E-07 | -0.159 | 0.027 | 4.94E-09 | -0.152 | 0.028 | 5.00E-08 | -0.139 | 0.027 | 2.69E-07 |
| *VAT1* | 0.139 | 0.024 | 5.85E-09 | 0.127 | 0.026 | 8.43E-07 | 0.117 | 0.026 | 6.81E-06 | 0.121 | 0.026 | 4.93E-06 | 0.113 | 0.026 | 9.47E-06 |
| *PAFAH1B3* | 0.144 | 0.025 | 1.20E-08 | 0.105 | 0.027 | 1.05E-04 | 0.103 | 0.027 | 1.54E-04 | 0.100 | 0.028 | 3.34E-04 | 0.106 | 0.027 | 7.58E-05 |
| *POLD1* | 0.158 | 0.028 | 1.57E-08 | 0.161 | 0.031 | 2.81E-07 | 0.164 | 0.031 | 1.79E-07 | 0.161 | 0.032 | 4.88E-07 | 0.153 | 0.031 | 6.68E-07 |
| *IVD* | -0.106 | 0.019 | 2.13E-08 | -0.094 | 0.024 | 6.36E-05 | -0.109 | 0.024 | 5.76E-06 | -0.094 | 0.024 | 1.10E-04 | -0.118 | 0.024 | 5.85E-07 |
| *ARG2* | 0.275 | 0.050 | 2.82E-08 | 0.236 | 0.053 | 9.86E-06 | 0.270 | 0.054 | 5.05E-07 | 0.253 | 0.055 | 3.66E-06 | 0.257 | 0.054 | 1.47E-06 |
| *SLC6A9* | 0.239 | 0.043 | 2.93E-08 | 0.225 | 0.044 | 2.79E-07 | 0.230 | 0.044 | 1.70E-07 | 0.219 | 0.045 | 1.05E-06 | 0.229 | 0.043 | 1.30E-07 |
| *TMCC2* | 0.149 | 0.027 | 5.06E-08 | 0.131 | 0.029 | 5.51E-06 | 0.148 | 0.029 | 4.09E-07 | 0.134 | 0.030 | 6.86E-06 | 0.139 | 0.029 | 1.28E-06 |
| *CLIP2* | 0.135 | 0.025 | 6.01E-08 | 0.132 | 0.026 | 6.04E-07 | 0.133 | 0.027 | 5.09E-07 | 0.125 | 0.027 | 3.47E-06 | 0.140 | 0.026 | 8.72E-08 |
| *ELP2* | -0.067 | 0.013 | 7.27E-08 | -0.047 | 0.013 | 2.87E-04 | -0.048 | 0.013 | 2.17E-04 | -0.049 | 0.013 | 2.53E-04 | -0.046 | 0.013 | 3.97E-04 |
| *ARSG* | -0.146 | 0.027 | 7.80E-08 | -0.118 | 0.031 | 1.46E-04 | -0.113 | 0.031 | 2.77E-04 | -0.113 | 0.032 | 3.71E-04 | -0.108 | 0.031 | 4.29E-04 |
| *QDPR* | 0.224 | 0.042 | 1.02E-07 | 0.193 | 0.043 | 6.51E-06 | 0.198 | 0.043 | 4.13E-06 | 0.177 | 0.044 | 5.28E-05 | 0.211 | 0.043 | 7.81E-07 |
| *NSMCE1* | -0.103 | 0.019 | 1.03E-07 | -0.114 | 0.023 | 7.84E-07 | -0.117 | 0.023 | 4.22E-07 | -0.106 | 0.024 | 6.66E-06 | -0.134 | 0.023 | 5.57E-09 |
| *IL2RG* | 0.394 | 0.074 | 1.04E-07 | 0.401 | 0.080 | 4.63E-07 | 0.403 | 0.079 | 3.83E-07 | 0.415 | 0.081 | 3.16E-07 | 0.364 | 0.079 | 3.52E-06 |
| *SCG3* | -0.166 | 0.032 | 1.34E-07 | -0.159 | 0.034 | 2.23E-06 | -0.166 | 0.034 | 7.84E-07 | -0.149 | 0.034 | 1.29E-05 | -0.181 | 0.034 | 6.81E-08 |
| *CXCR4* | 0.423 | 0.080 | 1.42E-07 | 0.322 | 0.082 | 7.89E-05 | 0.361 | 0.082 | 1.17E-05 | 0.306 | 0.083 | 2.40E-04 | 0.351 | 0.081 | 1.63E-05 |
| *RABGGTA* | 0.116 | 0.022 | 1.44E-07 | 0.089 | 0.024 | 2.17E-04 | 0.088 | 0.024 | 2.84E-04 | 0.079 | 0.025 | 1.30E-03 | 0.100 | 0.024 | 3.69E-05 |
| *RAB11FIP5* | 0.119 | 0.023 | 1.77E-07 | 0.098 | 0.026 | 2.10E-04 | 0.106 | 0.027 | 6.45E-05 | 0.100 | 0.027 | 2.25E-04 | 0.095 | 0.026 | 2.61E-04 |
| *HSD11B2* | 0.409 | 0.079 | 1.98E-07 | 0.361 | 0.086 | 2.46E-05 | 0.369 | 0.087 | 2.02E-05 | 0.336 | 0.088 | 1.37E-04 | 0.386 | 0.085 | 6.09E-06 |
| *MON1B* | -0.071 | 0.014 | 2.05E-07 | -0.053 | 0.015 | 4.25E-04 | -0.051 | 0.015 | 6.88E-04 | -0.052 | 0.015 | 8.50E-04 | -0.051 | 0.015 | 5.53E-04 |
| *PEPD* | 0.098 | 0.019 | 2.16E-07 | 0.078 | 0.028 | 4.53E-03 | 0.079 | 0.028 | 4.45E-03 | 0.079 | 0.029 | 5.45E-03 | 0.073 | 0.027 | 8.06E-03 |
| *SAP30L* | 0.129 | 0.025 | 2.25E-07 | 0.128 | 0.027 | 1.94E-06 | 0.127 | 0.027 | 2.68E-06 | 0.117 | 0.028 | 2.44E-05 | 0.136 | 0.027 | 3.95E-07 |
| *ZBBX* | -0.270 | 0.053 | 2.73E-07 | -0.245 | 0.057 | 1.69E-05 | -0.267 | 0.057 | 2.80E-06 | -0.255 | 0.058 | 1.19E-05 | -0.252 | 0.056 | 7.96E-06 |
| *USP31* | 0.149 | 0.029 | 2.85E-07 | 0.160 | 0.032 | 5.96E-07 | 0.153 | 0.032 | 2.25E-06 | 0.145 | 0.033 | 1.11E-05 | 0.167 | 0.032 | 1.70E-07 |
| *PRELP* | 0.225 | 0.044 | 2.90E-07 | 0.206 | 0.046 | 8.05E-06 | 0.215 | 0.047 | 4.86E-06 | 0.198 | 0.048 | 3.37E-05 | 0.226 | 0.046 | 1.08E-06 |
| *FHIP1B* | -0.121 | 0.024 | 2.95E-07 | -0.108 | 0.028 | 1.17E-04 | -0.124 | 0.028 | 1.21E-05 | -0.110 | 0.029 | 1.38E-04 | -0.125 | 0.028 | 7.02E-06 |
| *MARCHF8* | 0.113 | 0.022 | 3.17E-07 | 0.115 | 0.024 | 1.13E-06 | 0.123 | 0.024 | 2.10E-07 | 0.117 | 0.024 | 1.49E-06 | 0.115 | 0.024 | 1.05E-06 |
| *MED29* | -0.127 | 0.025 | 3.29E-07 | -0.126 | 0.030 | 1.98E-05 | -0.142 | 0.030 | 2.53E-06 | -0.125 | 0.030 | 3.86E-05 | -0.151 | 0.030 | 2.99E-07 |
| *SLC43A3* | 0.204 | 0.040 | 3.88E-07 | 0.186 | 0.042 | 8.57E-06 | 0.188 | 0.042 | 7.71E-06 | 0.182 | 0.043 | 2.23E-05 | 0.177 | 0.041 | 1.71E-05 |
| *ANP32E* | -0.130 | 0.026 | 4.26E-07 | -0.116 | 0.027 | 1.44E-05 | -0.129 | 0.027 | 2.43E-06 | -0.123 | 0.027 | 7.02E-06 | -0.119 | 0.027 | 8.89E-06 |
| *LRATD2* | 0.167 | 0.033 | 4.93E-07 | 0.183 | 0.037 | 7.34E-07 | 0.190 | 0.037 | 3.20E-07 | 0.176 | 0.038 | 3.54E-06 | 0.206 | 0.037 | 2.32E-08 |
| *PRR13* | -0.094 | 0.019 | 5.00E-07 | -0.089 | 0.024 | 1.79E-04 | -0.112 | 0.025 | 5.92E-06 | -0.093 | 0.025 | 1.68E-04 | -0.116 | 0.024 | 1.41E-06 |
| *PPM1D* | 0.131 | 0.026 | 5.55E-07 | 0.133 | 0.028 | 1.72E-06 | 0.131 | 0.028 | 2.69E-06 | 0.124 | 0.029 | 1.27E-05 | 0.140 | 0.028 | 3.82E-07 |
| *MTMR10* | 0.118 | 0.024 | 5.67E-07 | 0.117 | 0.024 | 1.44E-06 | 0.119 | 0.025 | 1.12E-06 | 0.113 | 0.025 | 6.23E-06 | 0.124 | 0.024 | 2.90E-07 |
| *WBP2NL* | -0.256 | 0.051 | 5.67E-07 | -0.187 | 0.053 | 4.17E-04 | -0.205 | 0.053 | 1.22E-04 | -0.184 | 0.055 | 7.36E-04 | -0.203 | 0.053 | 1.13E-04 |
| *STAG1* | 0.097 | 0.019 | 5.70E-07 | 0.111 | 0.024 | 4.28E-06 | 0.105 | 0.024 | 1.33E-05 | 0.100 | 0.025 | 5.22E-05 | 0.124 | 0.024 | 2.55E-07 |
| *GEM* | 0.379 | 0.076 | 5.92E-07 | 0.356 | 0.079 | 6.36E-06 | 0.372 | 0.080 | 2.87E-06 | 0.368 | 0.081 | 5.34E-06 | 0.363 | 0.079 | 4.13E-06 |
| *MYOT* | 0.265 | 0.053 | 6.06E-07 | 0.196 | 0.055 | 3.42E-04 | 0.202 | 0.056 | 2.71E-04 | 0.157 | 0.056 | 5.06E-03 | 0.259 | 0.056 | 3.14E-06 |
| *CHN2* | 0.187 | 0.038 | 6.53E-07 | 0.168 | 0.041 | 4.35E-05 | 0.173 | 0.041 | 2.94E-05 | 0.156 | 0.042 | 2.14E-04 | 0.184 | 0.041 | 7.29E-06 |
| *SMPX* | 0.252 | 0.051 | 6.99E-07 | 0.250 | 0.053 | 2.33E-06 | 0.236 | 0.053 | 7.89E-06 | 0.244 | 0.054 | 6.08E-06 | 0.214 | 0.052 | 4.03E-05 |
| *PRDM8* | 0.132 | 0.027 | 7.27E-07 | 0.146 | 0.031 | 1.80E-06 | 0.137 | 0.031 | 9.38E-06 | 0.132 | 0.032 | 3.16E-05 | 0.148 | 0.030 | 1.18E-06 |
| *EMP3* | 0.212 | 0.043 | 8.06E-07 | 0.178 | 0.047 | 1.54E-04 | 0.169 | 0.047 | 3.15E-04 | 0.179 | 0.048 | 1.89E-04 | 0.150 | 0.046 | 1.10E-03 |
| ***DYNLRB1*** | 0.062 | 0.013 | 8.63E-07 | 0.024 | 0.021 | 2.58E-01 | 0.022 | 0.021 | 2.98E-01 | 0.025 | 0.022 | 2.58E-01 | 0.018 | 0.021 | 3.80E-01 |
| *CNKSR3* | 0.172 | 0.035 | 9.63E-07 | 0.193 | 0.039 | 7.32E-07 | 0.204 | 0.039 | 1.49E-07 | 0.192 | 0.040 | 1.29E-06 | 0.201 | 0.039 | 1.88E-07 |
| *TNFRSF18* | -0.330 | 0.067 | 9.84E-07 | -0.341 | 0.069 | 6.97E-07 | -0.311 | 0.069 | 7.25E-06 | -0.317 | 0.071 | 7.49E-06 | -0.353 | 0.068 | 2.33E-07 |

**Supplementary Table 5:** Association results between cognitive domain scores and 13 DEGs in SymAD and AysmAD subjects across the ROSMAP, FHS and BUADRC datasets. Bold values indicate meta P-value<0.0038 and in at least two datasets were P<0.05.

| Domain | Gene | Meta-Analysis | | | | ROSMAP | | | | FHS | | | | BUADRC | | | |
|---|---|---|---|---|---|---|---|---|---|---|---|---|---|---|---|---|---|
| | | BETA | SE | Pvalue | Direction | BETA | SE | Tvalue | Pvalue | BETA | SE | Tvalue | Pvalue | BETA | SE | Tvalue | Pvalue |
| Memory | *ADAMTS2* | -0.0024 | 0.0004 | 1.45E-08 | --- | -0.00264 | 0.00046 | -5.71960 | 2.33E-08 | -0.00364 | 0.00467 | -0.77800 | 4.45E-01 | -0.00091 | 0.00126 | -0.72086 | 4.75E-01 |
| Memory | *SLC38A2* | -0.0002 | 0.0001 | 2.37E-05 | --- | -0.00030 | 0.00007 | -4.31913 | 2.05E-05 | -0.00010 | 0.00012 | -0.81595 | 4.23E-01 | -0.00016 | 0.00016 | -0.98943 | 3.28E-01 |
| Memory | *CHGA* | 0.0000 | 0.0000 | 3.56E-02 | -+- | -0.00004 | 0.00002 | -2.02299 | 4.38E-02 | 0.00003 | 0.00021 | 0.14334 | 8.87E-01 | -0.00005 | 0.00008 | -0.66104 | 5.12E-01 |
| Memory | *VAT1* | -0.0006 | 0.0002 | 4.27E-03 | --- | -0.00058 | 0.00026 | -2.20242 | 2.83E-02 | -0.00071 | 0.00069 | -1.03185 | 3.13E-01 | -0.00055 | 0.00036 | -1.51405 | 1.38E-01 |
| Memory | *PAFAH1B3* | -0.0015 | 0.0004 | 3.67E-04 | --- | -0.00158 | 0.00046 | -3.39998 | 7.53E-04 | -0.00319 | 0.00217 | -1.47280 | 1.55E-01 | -0.00053 | 0.00128 | -0.41631 | 6.79E-01 |
| Memory | *SLC6A9* | -0.0013 | 0.0003 | 1.56E-05 | --- | -0.00134 | 0.00036 | -3.71328 | 2.39E-04 | -0.00115 | 0.00076 | -1.50547 | 1.46E-01 | -0.00172 | 0.00103 | -1.67612 | 1.01E-01 |
| Memory | *TMCC2* | -0.0003 | 0.0001 | 3.82E-04 | --+ | -0.00036 | 0.00009 | -3.78854 | 1.79E-04 | -0.00069 | 0.00037 | -1.89413 | 7.14E-02 | 0.00014 | 0.00022 | 0.67029 | 5.06E-01 |
| Memory | *ARSG* | 0.0048 | 0.0012 | 1.12E-04 | +++ | 0.00476 | 0.00130 | 3.67678 | 2.74E-04 | 0.01153 | 0.00730 | 1.58121 | 1.28E-01 | 0.00172 | 0.00525 | 0.32766 | 7.45E-01 |
| Memory | *IL2RG* | -0.0056 | 0.0023 | 1.63E-02 | --- | -0.00488 | 0.00279 | -1.74886 | 8.12E-02 | -0.01595 | 0.00666 | -2.39607 | 2.55E-02 | -0.00137 | 0.00544 | -0.25117 | 8.03E-01 |
| Memory | *HSD11B2* | -0.0024 | 0.0007 | 8.03E-04 | --- | -0.00265 | 0.00079 | -3.36840 | 8.42E-04 | -0.00222 | 0.00706 | -0.31445 | 7.56E-01 | -0.00112 | 0.00184 | -0.60860 | 5.46E-01 |
| Memory | ***PRELP*** | -0.0004 | 0.0001 | 3.92E-05 | --+ | -0.00066 | 0.00014 | -4.65189 | 4.70E-06 | -0.00069 | 0.00025 | -2.71957 | 1.25E-02 | 0.00006 | 0.00017 | 0.34762 | 7.30E-01 |
| Memory | *FHIP1B* | 0.0012 | 0.0005 | 7.15E-03 | +-+ | 0.00194 | 0.00053 | 3.66685 | 2.84E-04 | -0.00177 | 0.00105 | -1.68123 | 1.07E-01 | 0.00234 | 0.00220 | 1.06397 | 2.94E-01 |
| Memory | ***EMP3*** | -0.0015 | 0.0005 | 7.75E-04 | --- | -0.00131 | 0.00057 | -2.32610 | 2.06E-02 | -0.00496 | 0.00157 | -3.15279 | 4.62E-03 | -0.00098 | 0.00088 | -1.11510 | 2.71E-01 |
| Language | *ADAMTS2* | -0.0025 | 0.0004 | 4.21E-09 | --- | -0.00264 | 0.00046 | -5.71609 | 2.42E-08 | -0.00590 | 0.00356 | -1.65776 | 1.14E-01 | -0.00127 | 0.00109 | -1.16661 | 2.50E-01 |
| Language | ***SLC38A2*** | -0.0002 | 0.0000 | 6.76E-05 | --- | -0.00022 | 0.00007 | -3.18947 | 1.56E-03 | -0.00008 | 0.00009 | -0.92850 | 3.65E-01 | -0.00038 | 0.00013 | -2.97893 | 4.96E-03 |
| Language | *CHGA* | -0.0001 | 0.0000 | 9.56E-03 | --- | -0.00005 | 0.00002 | -2.23353 | 2.62E-02 | -0.00003 | 0.00015 | -0.20972 | 8.36E-01 | -0.00010 | 0.00007 | -1.47993 | 1.47E-01 |
| Language | *VAT1* | -0.0004 | 0.0002 | 2.07E-02 | --- | -0.00034 | 0.00027 | -1.26983 | 2.05E-01 | -0.00055 | 0.00052 | -1.06805 | 2.99E-01 | -0.00054 | 0.00032 | -1.69837 | 9.74E-02 |
| Language | *PAFAH1B3* | -0.0015 | 0.0004 | 2.46E-04 | --- | -0.00151 | 0.00047 | -3.23038 | 1.36E-03 | -0.00177 | 0.00169 | -1.04626 | 3.09E-01 | -0.00152 | 0.00109 | -1.39198 | 1.72E-01 |
| Language | ***SLC6A9*** | -0.0009 | 0.0003 | 2.13E-03 | --- | -0.00086 | 0.00037 | -2.31933 | 2.10E-02 | -0.00056 | 0.00060 | -0.93791 | 3.60E-01 | -0.00195 | 0.00088 | -2.22123 | 3.22E-02 |
| Language | *TMCC2* | -0.0002 | 0.0001 | 1.99E-03 | --+ | -0.00030 | 0.00009 | -3.17179 | 1.66E-03 | -0.00034 | 0.00023 | -1.47864 | 1.56E-01 | 0.00004 | 0.00019 | 0.21654 | 8.30E-01 |
| Language | *ARSG* | 0.0038 | 0.0012 | 1.88E-03 | +++ | 0.00368 | 0.00129 | 2.84653 | 4.69E-03 | 0.00737 | 0.00617 | 1.19532 | 2.47E-01 | 0.00327 | 0.00466 | 0.70240 | 4.87E-01 |
| Language | *IL2RG* | -0.0023 | 0.0022 | 2.85E-01 | --- | -0.00261 | 0.00276 | -0.94662 | 3.45E-01 | -0.00188 | 0.00546 | -0.34352 | 7.35E-01 | -0.00187 | 0.00477 | -0.39282 | 6.97E-01 |
| Language | *HSD11B2* | -0.0028 | 0.0007 | 4.92E-05 | --- | -0.00292 | 0.00078 | -3.75360 | 2.06E-04 | -0.00832 | 0.00433 | -1.91939 | 7.01E-02 | -0.00156 | 0.00160 | -0.97388 | 3.36E-01 |
| Language | *PRELP* | -0.0003 | 0.0001 | 5.27E-03 | --- | -0.00055 | 0.00014 | -3.86443 | 1.34E-04 | -0.00003 | 0.00023 | -0.12708 | 9.00E-01 | -0.00005 | 0.00015 | -0.33749 | 7.38E-01 |
| Language | *FHIP1B* | 0.0009 | 0.0004 | 3.00E-02 | +-+ | 0.00148 | 0.00054 | 2.76986 | 5.92E-03 | -0.00040 | 0.00080 | -0.49941 | 6.23E-01 | 0.00178 | 0.00198 | 0.89998 | 3.74E-01 |
| Language | *EMP3* | -0.0009 | 0.0004 | 3.56E-02 | --- | -0.00080 | 0.00056 | -1.41259 | 1.59E-01 | -0.00167 | 0.00137 | -1.21378 | 2.40E-01 | -0.00087 | 0.00077 | -1.13793 | 2.62E-01 |
| Executive Function | ***ADAMTS2*** | -0.0021 | 0.0004 | 9.91E-09 | --- | -0.00219 | 0.00040 | -5.50561 | 7.69E-08 | -0.00624 | 0.00256 | -2.44205 | 2.52E-02 | -0.00079 | 0.00121 | -0.64928 | 5.22E-01 |
| Executive Function | *SLC38A2* | -0.0001 | 0.0000 | 2.32E-03 | --- | -0.00022 | 0.00006 | -3.75416 | 2.07E-04 | 0.00000 | 0.00007 | -0.01653 | 9.87E-01 | -0.00014 | 0.00016 | -0.88507 | 3.85E-01 |
| Executive Function | *CHGA* | 0.0000 | 0.0000 | 6.33E-03 | --- | -0.00005 | 0.00002 | -2.63721 | 8.78E-03 | -0.00007 | 0.00014 | -0.50776 | 6.18E-01 | -0.00004 | 0.00007 | -0.53152 | 6.00E-01 |
| Executive Function | *VAT1* | -0.0005 | 0.0002 | 7.97E-03 | --- | -0.00035 | 0.00023 | -1.52884 | 1.27E-01 | -0.00087 | 0.00041 | -2.13428 | 4.68E-02 | -0.00045 | 0.00037 | -1.19586 | 2.43E-01 |
| Executive Function | *PAFAH1B3* | -0.0013 | 0.0004 | 5.13E-04 | --- | -0.00115 | 0.00039 | -2.95573 | 3.36E-03 | -0.00115 | 0.00144 | -0.79945 | 4.34E-01 | -0.00236 | 0.00125 | -1.88254 | 7.19E-02 |
| Executive Function | *SLC6A9* | -0.0007 | 0.0003 | 1.06E-02 | --- | -0.00079 | 0.00032 | -2.41727 | 1.62E-02 | -0.00027 | 0.00048 | -0.55086 | 5.89E-01 | -0.00152 | 0.00114 | -1.32781 | 1.97E-01 |
| Executive Function | *TMCC2* | -0.0002 | 0.0001 | 1.72E-02 | --+ | -0.00025 | 0.00008 | -3.13603 | 1.88E-03 | -0.00019 | 0.00024 | -0.80476 | 4.31E-01 | 0.00034 | 0.00020 | 1.74883 | 9.31E-02 |
| Executive Function | ***ARSG*** | 0.0033 | 0.0010 | 1.09E-03 | ++- | 0.00323 | 0.00107 | 3.01524 | 2.78E-03 | 0.00992 | 0.00447 | 2.21894 | 3.96E-02 | -0.00228 | 0.00474 | -0.48034 | 6.35E-01 |
| Executive Function | *IL2RG* | -0.0038 | 0.0019 | 4.95E-02 | --- | -0.00372 | 0.00229 | -1.62537 | 1.05E-01 | -0.00279 | 0.00545 | -0.51139 | 6.15E-01 | -0.00472 | 0.00466 | -1.01418 | 3.21E-01 |
| Executive Function | *HSD11B2* | -0.0022 | 0.0006 | 3.67E-04 | --- | -0.00250 | 0.00067 | -3.71158 | 2.44E-04 | -0.00343 | 0.00439 | -0.78050 | 4.45E-01 | -0.00002 | 0.00183 | -0.01228 | 9.90E-01 |
| Executive Function | *PRELP* | -0.0003 | 0.0001 | 7.86E-04 | --+ | -0.00055 | 0.00012 | -4.57315 | 6.94E-06 | -0.00003 | 0.00027 | -0.11900 | 9.07E-01 | 0.00002 | 0.00015 | 0.10648 | 9.16E-01 |
| Executive Function | *FHIP1B* | 0.0011 | 0.0004 | 3.13E-03 | ++- | 0.00156 | 0.00045 | 3.50060 | 5.32E-04 | 0.00015 | 0.00066 | 0.22177 | 8.27E-01 | -0.00017 | 0.00190 | -0.08837 | 9.30E-01 |
| Executive Function | *EMP3* | -0.0011 | 0.0004 | 3.87E-03 | --- | -0.00078 | 0.00048 | -1.63818 | 1.02E-01 | -0.00189 | 0.00102 | -1.85258 | 8.04E-02 | -0.00201 | 0.00099 | -2.02632 | 5.40E-02 |

**Supplementary Table 6:** Association analysis between 13 DEGs and 12 neuropathological traits in SymAD and AsymAD subjects from the FHS and BUADRC datasets. Bold values indicate meta P-value<0.0038 and in at least two datasets were P<0.05.

| | | | Meta-Analysis | | | | FHS | | | | BUADRC | | | |
|---|---|---|---|---|---|---|---|---|---|---|---|---|---|---|
| Pathology | Trait | Gene | BETA | SE | Pvalue | Direction | BETA | SE | Tvalue | Pvalue | BETA | SE | Tvalue | Pvalue |
| Tau pathology | BraakStage | *ADAMTS2* | 0.0013 | 0.0010 | 1.75E-01 | ++ | 0.00296 | 0.00181 | 1.63562 | 1.08E-01 | 0.00065 | 0.00114 | 0.57403 | 5.70E-01 |
| Tau pathology | BraakStage | *SLC38A2* | 0.0002 | 0.0001 | 6.64E-03 | ++ | 0.00016 | 0.00006 | 2.76388 | 8.03E-03 | 0.00006 | 0.00016 | 0.34431 | 7.33E-01 |
| Tau pathology | BraakStage | *CHGA* | 0.0000 | 0.0001 | 9.63E-01 | -+ | -0.00007 | 0.00007 | -0.99993 | 3.22E-01 | 0.00007 | 0.00007 | 0.92951 | 3.59E-01 |
| Tau pathology | BraakStage | *VAT1* | 0.0007 | 0.0003 | 6.35E-03 | ++ | 0.00113 | 0.00037 | 3.01422 | 4.07E-03 | 0.00032 | 0.00034 | 0.93671 | 3.56E-01 |
| Tau pathology | BraakStage | *PAFAH1B3* | 0.0018 | 0.0008 | 3.39E-02 | ++ | 0.00149 | 0.00115 | 1.29754 | 2.01E-01 | 0.00211 | 0.00123 | 1.71795 | 9.52E-02 |
| Tau pathology | BraakStage | *SLC6A9* | 0.0010 | 0.0004 | 1.16E-02 | ++ | 0.00107 | 0.00044 | 2.41034 | 1.97E-02 | 0.00080 | 0.00100 | 0.79134 | 4.34E-01 |
| Tau pathology | BraakStage | *TMCC2* | 0.0001 | 0.0001 | 3.05E-01 | +- | 0.00037 | 0.00016 | 2.24861 | 2.91E-02 | -0.00025 | 0.00021 | -1.19911 | 2.39E-01 |
| Tau pathology | BraakStage | *ARSG* | -0.0010 | 0.0032 | 7.57E-01 | -- | -0.00009 | 0.00422 | -0.02217 | 9.82E-01 | -0.00209 | 0.00475 | -0.44058 | 6.62E-01 |
| Tau pathology | BraakStage | *IL2RG* | -0.0038 | 0.0036 | 3.00E-01 | -- | -0.00448 | 0.00487 | -0.92093 | 3.62E-01 | -0.00287 | 0.00545 | -0.52650 | 6.02E-01 |
| Tau pathology | BraakStage | *HSD11B2* | 0.0019 | 0.0015 | 1.89E-01 | ++ | 0.00279 | 0.00291 | 0.96142 | 3.41E-01 | 0.00164 | 0.00170 | 0.95966 | 3.44E-01 |
| Tau pathology | BraakStage | *PRELP* | 0.0000 | 0.0001 | 9.04E-01 | ++ | 0.00003 | 0.00019 | 0.15753 | 8.75E-01 | 0.00000 | 0.00015 | 0.02874 | 9.77E-01 |
| Tau pathology | BraakStage | *FHIP1B* | 0.0004 | 0.0005 | 4.77E-01 | +- | 0.00052 | 0.00057 | 0.91638 | 3.64E-01 | -0.00134 | 0.00206 | -0.64719 | 5.22E-01 |
| Tau pathology | BraakStage | *EMP3* | 0.0013 | 0.0007 | 4.97E-02 | ++ | 0.00209 | 0.00114 | 1.83483 | 7.26E-02 | 0.00090 | 0.00082 | 1.09862 | 2.80E-01 |
| Tau pathology | AT8 | *ADAMTS2* | 0.0012 | 0.0011 | 2.46E-01 | ++ | 0.00382 | 0.00248 | 1.54407 | 1.29E-01 | 0.00065 | 0.00117 | 0.55386 | 5.83E-01 |
| Tau pathology | AT8 | *SLC38A2* | 0.0001 | 0.0001 | 2.75E-01 | +- | 0.00011 | 0.00008 | 1.28783 | 2.04E-01 | -0.00001 | 0.00016 | -0.08393 | 9.34E-01 |
| Tau pathology | AT8 | *CHGA* | -0.0001 | 0.0001 | 1.99E-01 | -+ | -0.00023 | 0.00009 | -2.46402 | 1.74E-02 | 0.00004 | 0.00008 | 0.47007 | 6.41E-01 |
| Tau pathology | AT8 | *VAT1* | 0.0004 | 0.0003 | 1.43E-01 | ++ | 0.00105 | 0.00053 | 1.96513 | 5.52E-02 | 0.00016 | 0.00036 | 0.44371 | 6.60E-01 |
| Tau pathology | AT8 | *PAFAH1B3* | 0.0003 | 0.0010 | 7.25E-01 | -+ | -0.00273 | 0.00154 | -1.77200 | 8.27E-02 | 0.00254 | 0.00130 | 1.95715 | 5.73E-02 |
| Tau pathology | AT8 | *SLC6A9* | 0.0009 | 0.0005 | 8.87E-02 | +- | 0.00138 | 0.00061 | 2.28137 | 2.70E-02 | -0.00039 | 0.00095 | -0.40934 | 6.84E-01 |
| Tau pathology | AT8 | *TMCC2* | 0.0002 | 0.0002 | 2.46E-01 | -+ | -0.00006 | 0.00023 | -0.25215 | 8.02E-01 | 0.00037 | 0.00021 | 1.78872 | 8.12E-02 |
| Tau pathology | AT8 | ***ARSG*** | -0.0110 | 0.0035 | 1.50E-03 | -- | -0.01274 | 0.00545 | -2.33652 | 2.37E-02 | -0.00985 | 0.00450 | -2.18808 | 3.46E-02 |
| Tau pathology | AT8 | *IL2RG* | 0.0025 | 0.0040 | 5.41E-01 | +- | 0.00853 | 0.00657 | 1.29852 | 2.00E-01 | -0.00120 | 0.00510 | -0.23526 | 8.15E-01 |
| Tau pathology | AT8 | *HSD11B2* | 0.0017 | 0.0016 | 2.77E-01 | ++ | 0.00125 | 0.00404 | 0.30983 | 7.58E-01 | 0.00184 | 0.00175 | 1.04959 | 3.00E-01 |
| Tau pathology | AT8 | *PRELP* | 0.0003 | 0.0001 | 5.41E-02 | ++ | 0.00060 | 0.00024 | 2.47334 | 1.70E-02 | 0.00011 | 0.00015 | 0.70434 | 4.85E-01 |
| Tau pathology | AT8 | *FHIP1B* | 0.0013 | 0.0007 | 7.03E-02 | +- | 0.00174 | 0.00074 | 2.36512 | 2.21E-02 | -0.00273 | 0.00212 | -1.28929 | 2.05E-01 |
| Tau pathology | AT8 | *EMP3* | 0.0008 | 0.0008 | 2.88E-01 | ++ | 0.00278 | 0.00156 | 1.78761 | 8.02E-02 | 0.00019 | 0.00087 | 0.21798 | 8.29E-01 |
| Tau pathology | pTau181 | *ADAMTS2* | 0.0016 | 0.0010 | 1.01E-01 | ++ | 0.00683 | 0.00317 | 2.15239 | 3.63E-02 | 0.00104 | 0.00100 | 1.03880 | 3.05E-01 |
| Tau pathology | pTau181 | *SLC38A2* | 0.0000 | 0.0001 | 5.89E-01 | +- | 0.00016 | 0.00011 | 1.41665 | 1.63E-01 | -0.00013 | 0.00014 | -0.91537 | 3.65E-01 |
| Tau pathology | pTau181 | *CHGA* | 0.0000 | 0.0001 | 8.58E-01 | -+ | -0.00028 | 0.00012 | -2.28177 | 2.69E-02 | 0.00007 | 0.00007 | 1.06315 | 2.94E-01 |
| Tau pathology | pTau181 | *VAT1* | 0.0002 | 0.0003 | 5.87E-01 | +- | 0.00166 | 0.00069 | 2.41116 | 1.97E-02 | -0.00018 | 0.00033 | -0.54587 | 5.88E-01 |
| Tau pathology | pTau181 | *PAFAH1B3* | -0.0001 | 0.0010 | 8.87E-01 | -+ | -0.00208 | 0.00206 | -1.00997 | 3.17E-01 | 0.00046 | 0.00115 | 0.40208 | 6.90E-01 |
| Tau pathology | pTau181 | *SLC6A9* | 0.0018 | 0.0006 | 1.50E-03 | ++ | 0.00241 | 0.00076 | 3.16760 | 2.65E-03 | 0.00104 | 0.00085 | 1.22228 | 2.28E-01 |
| Tau pathology | pTau181 | *TMCC2* | -0.0001 | 0.0002 | 7.49E-01 | +- | 0.00032 | 0.00030 | 1.05252 | 2.98E-01 | -0.00020 | 0.00019 | -1.04993 | 2.99E-01 |
| Tau pathology | pTau181 | *ARSG* | -0.0005 | 0.0038 | 8.91E-01 | +- | 0.00198 | 0.00754 | 0.26274 | 7.94E-01 | -0.00138 | 0.00441 | -0.31333 | 7.56E-01 |
| Tau pathology | pTau181 | *IL2RG* | -0.0026 | 0.0042 | 5.38E-01 | -- | -0.00394 | 0.00875 | -0.45070 | 6.54E-01 | -0.00217 | 0.00476 | -0.45529 | 6.51E-01 |
| Tau pathology | pTau181 | *HSD11B2* | 0.0020 | 0.0015 | 1.71E-01 | ++ | 0.00793 | 0.00511 | 1.55032 | 1.28E-01 | 0.00148 | 0.00153 | 0.96423 | 3.40E-01 |
| Tau pathology | pTau181 | *PRELP* | -0.0001 | 0.0001 | 4.19E-01 | +- | 0.00038 | 0.00033 | 1.14084 | 2.59E-01 | -0.00019 | 0.00014 | -1.36971 | 1.78E-01 |
| Tau pathology | pTau181 | *FHIP1B* | 0.0025 | 0.0009 | 3.64E-03 | ++ | 0.00258 | 0.00095 | 2.70421 | 9.39E-03 | 0.00208 | 0.00190 | 1.09540 | 2.79E-01 |
| Tau pathology | pTau181 | *EMP3* | 0.0004 | 0.0007 | 6.37E-01 | ++ | 0.00158 | 0.00209 | 0.75650 | 4.53E-01 | 0.00017 | 0.00080 | 0.21593 | 8.30E-01 |
| Tau pathology | pTau231 | *ADAMTS2* | -0.0004 | 0.0010 | 7.05E-01 | +- | 0.00974 | 0.00318 | 3.06284 | 3.56E-03 | -0.00162 | 0.00110 | -1.46479 | 1.50E-01 |
| Tau pathology | pTau231 | *SLC38A2* | 0.0000 | 0.0001 | 9.05E-01 | +- | 0.00022 | 0.00011 | 1.96723 | 5.48E-02 | -0.00034 | 0.00015 | -2.33705 | 2.41E-02 |
| Tau pathology | pTau231 | *CHGA* | -0.0001 | 0.0001 | 4.42E-01 | -- | -0.00012 | 0.00013 | -0.89791 | 3.74E-01 | -0.00003 | 0.00008 | -0.37362 | 7.10E-01 |
| Tau pathology | pTau231 | *VAT1* | 0.0005 | 0.0003 | 1.37E-01 | +- | 0.00278 | 0.00065 | 4.26586 | 9.07E-05 | -0.00024 | 0.00036 | -0.66813 | 5.08E-01 |
| Tau pathology | pTau231 | *PAFAH1B3* | -0.0015 | 0.0011 | 1.53E-01 | -- | -0.00300 | 0.00214 | -1.40489 | 1.66E-01 | -0.00104 | 0.00124 | -0.83408 | 4.09E-01 |
| Tau pathology | pTau231 | *SLC6A9* | 0.0017 | 0.0006 | 6.04E-03 | ++ | 0.00276 | 0.00078 | 3.54143 | 8.84E-04 | 0.00000 | 0.00096 | 0.00002 | 1.00E+00 |
| Tau pathology | pTau231 | *TMCC2* | 0.0001 | 0.0002 | 5.98E-01 | +- | 0.00043 | 0.00031 | 1.37590 | 1.75E-01 | -0.00007 | 0.00021 | -0.30400 | 7.63E-01 |
| Tau pathology | pTau231 | *ARSG* | 0.0061 | 0.0040 | 1.32E-01 | ++ | 0.00231 | 0.00788 | 0.29350 | 7.70E-01 | 0.00738 | 0.00468 | 1.57809 | 1.22E-01 |
| Tau pathology | pTau231 | *IL2RG* | -0.0010 | 0.0046 | 8.25E-01 | -- | -0.00334 | 0.00916 | -0.36523 | 7.17E-01 | -0.00023 | 0.00527 | -0.04451 | 9.65E-01 |
| Tau pathology | pTau231 | *HSD11B2* | -0.0002 | 0.0016 | 9.22E-01 | +- | 0.00969 | 0.00530 | 1.82881 | 7.35E-02 | -0.00118 | 0.00171 | -0.69242 | 4.92E-01 |
| Tau pathology | pTau231 | *PRELP* | -0.0001 | 0.0001 | 4.54E-01 | +- | 0.00026 | 0.00035 | 0.75564 | 4.53E-01 | -0.00018 | 0.00016 | -1.16125 | 2.52E-01 |
| Tau pathology | pTau231 | *FHIP1B* | 0.0026 | 0.0009 | 4.99E-03 | ++ | 0.00238 | 0.00101 | 2.34621 | 2.31E-02 | 0.00328 | 0.00206 | 1.59113 | 1.19E-01 |
| Tau pathology | pTau231 | *EMP3* | 0.0007 | 0.0008 | 3.73E-01 | ++ | 0.00252 | 0.00217 | 1.15951 | 2.52E-01 | 0.00043 | 0.00089 | 0.48859 | 6.28E-01 |
| Tau pathology | pTau202 | *ADAMTS2* | 0.0006 | 0.0011 | 6.09E-01 | ++ | 0.00322 | 0.00337 | 0.95632 | 3.44E-01 | 0.00024 | 0.00120 | 0.20075 | 8.42E-01 |
| Tau pathology | pTau202 | *SLC38A2* | 0.0002 | 0.0001 | 9.87E-02 | ++ | 0.00020 | 0.00011 | 1.72872 | 9.03E-02 | 0.00007 | 0.00017 | 0.40214 | 6.90E-01 |
| Tau pathology | pTau202 | *CHGA* | -0.0001 | 0.0001 | 3.91E-01 | -- | -0.00006 | 0.00013 | -0.46079 | 6.47E-01 | -0.00006 | 0.00008 | -0.72378 | 4.73E-01 |
| Tau pathology | pTau202 | *VAT1* | 0.0001 | 0.0003 | 7.28E-01 | +- | 0.00177 | 0.00072 | 2.45006 | 1.80E-02 | -0.00035 | 0.00039 | -0.91172 | 3.67E-01 |
| Tau pathology | pTau202 | *PAFAH1B3* | -0.0001 | 0.0011 | 9.54E-01 | -+ | -0.00257 | 0.00213 | -1.20338 | 2.35E-01 | 0.00089 | 0.00132 | 0.67686 | 5.02E-01 |
| Tau pathology | pTau202 | *SLC6A9* | 0.0011 | 0.0006 | 6.85E-02 | +- | 0.00218 | 0.00080 | 2.72107 | 9.04E-03 | -0.00050 | 0.00101 | -0.49891 | 6.20E-01 |
| Tau pathology | pTau202 | *TMCC2* | 0.0002 | 0.0002 | 3.60E-01 | +- | 0.00076 | 0.00029 | 2.58257 | 1.29E-02 | -0.00019 | 0.00023 | -0.85125 | 3.99E-01 |
| Tau pathology | pTau202 | *ARSG* | -0.0002 | 0.0042 | 9.57E-01 | +- | 0.01390 | 0.00751 | 1.85212 | 7.02E-02 | -0.00655 | 0.00502 | -1.30482 | 1.99E-01 |
| Tau pathology | pTau202 | *IL2RG* | -0.0069 | 0.0049 | 1.57E-01 | -- | -0.01971 | 0.01063 | -1.85451 | 6.98E-02 | -0.00349 | 0.00553 | -0.63072 | 5.32E-01 |
| Tau pathology | pTau202 | *HSD11B2* | 0.0005 | 0.0017 | 7.62E-01 | +- | 0.00598 | 0.00529 | 1.13084 | 2.64E-01 | -0.00012 | 0.00182 | -0.06812 | 9.46E-01 |
| Tau pathology | pTau202 | *PRELP* | 0.0000 | 0.0002 | 7.70E-01 | ++ | 0.00026 | 0.00037 | 0.69548 | 4.90E-01 | 0.00000 | 0.00017 | 0.00851 | 9.93E-01 |
| Tau pathology | pTau202 | *FHIP1B* | 0.0020 | 0.0009 | 2.41E-02 | +- | 0.00264 | 0.00098 | 2.69496 | 9.67E-03 | -0.00125 | 0.00227 | -0.55030 | 5.85E-01 |
| Tau pathology | pTau202 | *EMP3* | 0.0009 | 0.0009 | 3.38E-01 | ++ | 0.00082 | 0.00216 | 0.37937 | 7.06E-01 | 0.00086 | 0.00098 | 0.88031 | 3.84E-01 |
| Tau pathology | pTau396 | *ADAMTS2* | 0.0022 | 0.0011 | 3.60E-02 | ++ | 0.00361 | 0.00319 | 1.13160 | 2.63E-01 | 0.00207 | 0.00113 | 1.82396 | 7.53E-02 |
| Tau pathology | pTau396 | *SLC38A2* | 0.0002 | 0.0001 | 8.39E-02 | ++ | 0.00020 | 0.00011 | 1.87136 | 6.74E-02 | 0.00005 | 0.00016 | 0.31026 | 7.58E-01 |
| Tau pathology | pTau396 | *CHGA* | 0.0000 | 0.0001 | 9.82E-01 | -+ | -0.00003 | 0.00013 | -0.22076 | 8.26E-01 | 0.00001 | 0.00008 | 0.11331 | 9.10E-01 |
| Tau pathology | pTau396 | *VAT1* | 0.0006 | 0.0003 | 4.96E-02 | ++ | 0.00188 | 0.00068 | 2.76369 | 8.08E-03 | 0.00027 | 0.00038 | 0.70508 | 4.85E-01 |
| Tau pathology | pTau396 | *PAFAH1B3* | 0.0008 | 0.0011 | 4.41E-01 | -+ | -0.00363 | 0.00199 | -1.82289 | 7.46E-02 | 0.00253 | 0.00124 | 2.03979 | 4.77E-02 |
| Tau pathology | pTau396 | *SLC6A9* | 0.0018 | 0.0006 | 2.05E-03 | ++ | 0.00241 | 0.00074 | 3.25420 | 2.09E-03 | 0.00079 | 0.00098 | 0.80448 | 4.26E-01 |
| Tau pathology | pTau396 | *TMCC2* | 0.0004 | 0.0002 | 2.43E-02 | ++ | 0.00082 | 0.00027 | 3.01350 | 4.12E-03 | 0.00010 | 0.00022 | 0.43617 | 6.65E-01 |
| Tau pathology | pTau396 | *ARSG* | -0.0025 | 0.0039 | 5.19E-01 | +- | 0.01596 | 0.00703 | 2.27044 | 2.77E-02 | -0.01088 | 0.00473 | -2.30231 | 2.63E-02 |
| Tau pathology | pTau396 | *IL2RG* | -0.0072 | 0.0048 | 1.33E-01 | -- | -0.01458 | 0.01027 | -1.42025 | 1.62E-01 | -0.00513 | 0.00538 | -0.95285 | 3.46E-01 |
| Tau pathology | pTau396 | *HSD11B2* | 0.0030 | 0.0016 | 6.69E-02 | ++ | 0.00538 | 0.00504 | 1.06576 | 2.92E-01 | 0.00272 | 0.00173 | 1.57139 | 1.24E-01 |
| Tau pathology | pTau396 | *PRELP* | 0.0001 | 0.0001 | 4.37E-01 | ++ | 0.00012 | 0.00036 | 0.33842 | 7.37E-01 | 0.00011 | 0.00016 | 0.69981 | 4.88E-01 |
| Tau pathology | pTau396 | *FHIP1B* | 0.0023 | 0.0008 | 6.27E-03 | +- | 0.00298 | 0.00090 | 3.29488 | 1.86E-03 | -0.00184 | 0.00221 | -0.83346 | 4.09E-01 |
| Tau pathology | pTau396 | *EMP3* | 0.0010 | 0.0009 | 2.24E-01 | -+ | -0.00141 | 0.00205 | -0.68491 | 4.97E-01 | 0.00154 | 0.00093 | 1.64935 | 1.07E-01 |

| | | | | | | | | | | | | | | |
|---|---|---|---|---|---|---|---|---|---|---|---|---|---|---|
| Beta-amyloid pathology | CERAD | *ADAMTS2* | 0.0000 | 0.0007 | 9.62E-01 | +- | 0.00170 | 0.00204 | 0.83250 | 4.09E-01 | -0.00028 | 0.00077 | -0.36570 | 7.17E-01 |
| Beta-amyloid pathology | CERAD | *SLC38A2* | 0.0000 | 0.0001 | 4.86E-01 | +- | 0.00006 | 0.00007 | 0.88072 | 3.83E-01 | -0.00001 | 0.00011 | -0.08158 | 9.36E-01 |
| Beta-amyloid pathology | CERAD | *CHGA* | 0.0000 | 0.0000 | 8.14E-01 | -+ | 0.00000 | 0.00008 | -0.06262 | 9.50E-01 | 0.00002 | 0.00005 | 0.31269 | 7.57E-01 |
| Beta-amyloid pathology | CERAD | *VAT1* | 0.0001 | 0.0002 | 6.04E-01 | +- | 0.00098 | 0.00043 | 2.28536 | 2.67E-02 | -0.00016 | 0.00024 | -0.66976 | 5.09E-01 |
| Beta-amyloid pathology | CERAD | *PAFAH1B3* | 0.0001 | 0.0007 | 9.13E-01 | +- | 0.00336 | 0.00120 | 2.79984 | 7.29E-03 | -0.00163 | 0.00086 | -1.88368 | 7.04E-02 |
| Beta-amyloid pathology | CERAD | *SLC6A9* | 0.0004 | 0.0004 | 3.74E-01 | ++ | 0.00049 | 0.00051 | 0.94544 | 3.49E-01 | 0.00015 | 0.00069 | 0.21745 | 8.29E-01 |
| Beta-amyloid pathology | CERAD | *TMCC2* | 0.0001 | 0.0001 | 5.49E-01 | +- | 0.00029 | 0.00018 | 1.59957 | 1.16E-01 | -0.00007 | 0.00014 | -0.48940 | 6.29E-01 |
| Beta-amyloid pathology | CERAD | *ARSG* | -0.0019 | 0.0027 | 4.95E-01 | +- | 0.00113 | 0.00468 | 0.24116 | 8.10E-01 | -0.00338 | 0.00335 | -1.01140 | 3.21E-01 |
| Beta-amyloid pathology | CERAD | *IL2RG* | -0.0016 | 0.0033 | 6.34E-01 | -- | -0.00153 | 0.00544 | -0.28094 | 7.80E-01 | -0.00163 | 0.00424 | -0.38547 | 7.03E-01 |
| Beta-amyloid pathology | CERAD | *HSD11B2* | 0.0002 | 0.0011 | 8.35E-01 | +- | 0.00217 | 0.00323 | 0.66953 | 5.06E-01 | -0.00003 | 0.00117 | -0.02129 | 9.83E-01 |
| Beta-amyloid pathology | CERAD | *PRELP* | -0.0001 | 0.0001 | 4.25E-01 | -- | -0.00009 | 0.00021 | -0.43919 | 6.62E-01 | -0.00007 | 0.00010 | -0.67445 | 5.06E-01 |
| Beta-amyloid pathology | CERAD | *FHIP1B* | 0.0000 | 0.0006 | 9.97E-01 | -+ | -0.00037 | 0.00063 | -0.57951 | 5.65E-01 | 0.00169 | 0.00136 | 1.24089 | 2.25E-01 |
| Beta-amyloid pathology | CERAD | *EMP3* | 0.0000 | 0.0005 | 9.94E-01 | +- | 0.00292 | 0.00124 | 2.35839 | 2.24E-02 | -0.00058 | 0.00055 | -1.04690 | 3.04E-01 |
| Beta-amyloid pathology | Ab40 | *ADAMTS2* | -0.0009 | 0.0011 | 3.74E-01 | +- | 0.00036 | 0.00311 | 0.11463 | 9.09E-01 | -0.00112 | 0.00114 | -0.98767 | 3.29E-01 |
| Beta-amyloid pathology | Ab40 | *SLC38A2* | -0.0001 | 0.0001 | 4.27E-01 | -- | -0.00002 | 0.00011 | -0.17904 | 8.59E-01 | -0.00018 | 0.00016 | -1.14891 | 2.57E-01 |
| Beta-amyloid pathology | Ab40 | *CHGA* | 0.0000 | 0.0001 | 9.54E-01 | -+ | -0.00003 | 0.00012 | -0.24211 | 8.10E-01 | 0.00001 | 0.00008 | 0.08612 | 9.32E-01 |
| Beta-amyloid pathology | Ab40 | *VAT1* | 0.0006 | 0.0003 | 4.11E-02 | -+ | -0.00016 | 0.00068 | -0.23069 | 8.19E-01 | 0.00084 | 0.00035 | 2.40861 | 2.03E-02 |
| Beta-amyloid pathology | Ab40 | *PAFAH1B3* | 0.0015 | 0.0011 | 1.65E-01 | ++ | 0.00219 | 0.00193 | 1.13675 | 2.61E-01 | 0.00115 | 0.00126 | 0.91383 | 3.66E-01 |
| Beta-amyloid pathology | Ab40 | *SLC6A9* | 0.0006 | 0.0006 | 3.26E-01 | ++ | 0.00011 | 0.00078 | 0.13492 | 8.93E-01 | 0.00132 | 0.00095 | 1.38228 | 1.74E-01 |
| Beta-amyloid pathology | Ab40 | *TMCC2* | 0.0001 | 0.0002 | 4.05E-01 | ++ | 0.00008 | 0.00029 | 0.27860 | 7.82E-01 | 0.00018 | 0.00022 | 0.83412 | 4.09E-01 |
| Beta-amyloid pathology | Ab40 | *ARSG* | 0.0049 | 0.0040 | 2.22E-01 | ++ | 0.00298 | 0.00707 | 0.42113 | 6.76E-01 | 0.00574 | 0.00482 | 1.19260 | 2.39E-01 |
| Beta-amyloid pathology | Ab40 | *IL2RG* | 0.0087 | 0.0043 | 4.15E-02 | -+ | -0.00141 | 0.00823 | -0.17155 | 8.64E-01 | 0.01250 | 0.00501 | 2.49226 | 1.65E-02 |
| Beta-amyloid pathology | Ab40 | *HSD11B2* | -0.0013 | 0.0016 | 4.19E-01 | -- | -0.00141 | 0.00491 | -0.28679 | 7.75E-01 | -0.00131 | 0.00174 | -0.75578 | 4.54E-01 |
| Beta-amyloid pathology | Ab40 | *PRELP* | 0.0000 | 0.0001 | 9.54E-01 | -+ | -0.00007 | 0.00031 | -0.22808 | 8.21E-01 | 0.00003 | 0.00016 | 0.18235 | 8.56E-01 |
| Beta-amyloid pathology | Ab40 | *FHIP1B* | -0.0003 | 0.0009 | 7.72E-01 | -+ | -0.00044 | 0.00096 | -0.45891 | 6.48E-01 | 0.00068 | 0.00215 | 0.31739 | 7.52E-01 |
| Beta-amyloid pathology | Ab40 | *EMP3* | 0.0012 | 0.0008 | 1.41E-01 | ++ | 0.00175 | 0.00196 | 0.88938 | 3.78E-01 | 0.00108 | 0.00089 | 1.21543 | 2.31E-01 |
| Beta-amyloid pathology | Ab42 | *ADAMTS2* | -0.0014 | 0.0009 | 1.25E-01 | -- | -0.00482 | 0.00258 | -1.86493 | 6.82E-02 | -0.00091 | 0.00097 | -0.93689 | 3.54E-01 |
| Beta-amyloid pathology | Ab42 | *SLC38A2* | -0.0001 | 0.0001 | 1.08E-01 | -- | -0.00016 | 0.00009 | -1.85930 | 6.90E-02 | -0.00001 | 0.00013 | -0.09392 | 9.26E-01 |
| Beta-amyloid pathology | Ab42 | *CHGA* | 0.0000 | 0.0001 | 9.15E-01 | +- | 0.00010 | 0.00010 | 0.94602 | 3.49E-01 | -0.00003 | 0.00007 | -0.47896 | 6.34E-01 |
| Beta-amyloid pathology | Ab42 | *VAT1* | 0.0000 | 0.0003 | 8.60E-01 | -+ | -0.00119 | 0.00056 | -2.10968 | 4.00E-02 | 0.00042 | 0.00031 | 1.35919 | 1.81E-01 |
| Beta-amyloid pathology | Ab42 | *PAFAH1B3* | 0.0016 | 0.0009 | 7.14E-02 | ++ | 0.00272 | 0.00163 | 1.66554 | 1.02E-01 | 0.00114 | 0.00107 | 1.06308 | 2.94E-01 |
| Beta-amyloid pathology | Ab42 | *SLC6A9* | -0.0007 | 0.0005 | 1.96E-01 | -- | -0.00087 | 0.00066 | -1.30643 | 1.98E-01 | -0.00036 | 0.00083 | -0.43647 | 6.65E-01 |
| Beta-amyloid pathology | Ab42 | *TMCC2* | -0.0002 | 0.0001 | 1.87E-01 | -- | -0.00020 | 0.00024 | -0.83860 | 4.06E-01 | -0.00019 | 0.00018 | -1.01967 | 3.13E-01 |
| Beta-amyloid pathology | Ab42 | *ARSG* | -0.0014 | 0.0034 | 6.84E-01 | -- | -0.00247 | 0.00607 | -0.40762 | 6.85E-01 | -0.00089 | 0.00417 | -0.21332 | 8.32E-01 |
| Beta-amyloid pathology | Ab42 | *IL2RG* | -0.0054 | 0.0038 | 1.56E-01 | -- | -0.00938 | 0.00694 | -1.35234 | 1.82E-01 | -0.00368 | 0.00453 | -0.81234 | 4.21E-01 |
| Beta-amyloid pathology | Ab42 | *HSD11B2* | -0.0015 | 0.0014 | 2.66E-01 | -- | -0.00216 | 0.00421 | -0.51285 | 6.10E-01 | -0.00147 | 0.00147 | -0.99798 | 3.24E-01 |
| Beta-amyloid pathology | Ab42 | *PRELP* | 0.0000 | 0.0001 | 8.57E-01 | -+ | -0.00028 | 0.00027 | -1.06848 | 2.91E-01 | 0.00005 | 0.00014 | 0.34933 | 7.29E-01 |
| Beta-amyloid pathology | Ab42 | *FHIP1B* | -0.0010 | 0.0007 | 1.46E-01 | -+ | -0.00181 | 0.00078 | -2.31313 | 2.50E-02 | 0.00294 | 0.00178 | 1.64754 | 1.07E-01 |
| Beta-amyloid pathology | Ab42 | *EMP3* | 0.0005 | 0.0007 | 4.75E-01 | -+ | -0.00069 | 0.00170 | -0.40889 | 6.84E-01 | 0.00074 | 0.00076 | 0.96728 | 3.39E-01 |
| Synaptic density | PSD.95 | *ADAMTS2* | -0.0009 | 0.0011 | 4.27E-01 | -- | -0.00564 | 0.00302 | -1.86627 | 6.80E-02 | -0.00016 | 0.00116 | -0.13618 | 8.92E-01 |
| Synaptic density | PSD.95 | *SLC38A2* | -0.0001 | 0.0001 | 5.53E-01 | -- | -0.00003 | 0.00011 | -0.31602 | 7.53E-01 | -0.00009 | 0.00016 | -0.59298 | 5.56E-01 |
| Synaptic density | PSD.95 | *CHGA* | 0.0000 | 0.0001 | 9.33E-01 | -+ | -0.00014 | 0.00012 | -1.21139 | 2.32E-01 | 0.00005 | 0.00008 | 0.68378 | 4.98E-01 |
| Synaptic density | PSD.95 | *VAT1* | -0.0008 | 0.0003 | 9.56E-03 | +- | 0.00083 | 0.00068 | 1.23060 | 2.24E-01 | -0.00115 | 0.00033 | -3.48011 | 1.14E-03 |
| Synaptic density | PSD.95 | *PAFAH1B3* | -0.0009 | 0.0011 | 3.84E-01 | -- | -0.00215 | 0.00194 | -1.10705 | 2.74E-01 | -0.00040 | 0.00128 | -0.31279 | 7.56E-01 |
| Synaptic density | PSD.95 | *SLC6A9* | 0.0001 | 0.0006 | 8.39E-01 | ++ | 0.00009 | 0.00079 | 0.11431 | 9.09E-01 | 0.00018 | 0.00098 | 0.18126 | 8.57E-01 |
| Synaptic density | PSD.95 | *TMCC2* | -0.0004 | 0.0002 | 3.31E-02 | -- | -0.00015 | 0.00029 | -0.53156 | 5.97E-01 | -0.00047 | 0.00021 | -2.24637 | 2.97E-02 |
| Synaptic density | PSD.95 | *ARSG* | -0.0028 | 0.0040 | 4.90E-01 | -- | -0.00062 | 0.00711 | -0.08674 | 9.31E-01 | -0.00380 | 0.00489 | -0.77877 | 4.40E-01 |
| Synaptic density | PSD.95 | *IL2RG* | -0.0023 | 0.0045 | 6.13E-01 | +- | 0.00357 | 0.00825 | 0.43296 | 6.67E-01 | -0.00471 | 0.00534 | -0.88295 | 3.82E-01 |
| Synaptic density | PSD.95 | *HSD11B2* | -0.0010 | 0.0016 | 5.55E-01 | -+ | -0.00925 | 0.00476 | -1.94364 | 5.77E-02 | 0.00016 | 0.00176 | 0.08923 | 9.29E-01 |
| Synaptic density | PSD.95 | *PRELP* | -0.0005 | 0.0001 | 6.18E-06 | +- | 0.00029 | 0.00031 | 0.91490 | 3.65E-01 | -0.00067 | 0.00013 | -5.25488 | 4.14E-06 |
| Synaptic density | PSD.95 | *FHIP1B* | 0.0014 | 0.0009 | 1.10E-01 | ++ | 0.00124 | 0.00095 | 1.30357 | 1.98E-01 | 0.00215 | 0.00214 | 1.00312 | 3.21E-01 |
| Synaptic density | PSD.95 | *EMP3* | -0.0001 | 0.0008 | 8.95E-01 | +- | 0.00310 | 0.00194 | 1.60288 | 1.15E-01 | -0.00080 | 0.00090 | -0.89070 | 3.78E-01 |
| Synaptic density | Asyn | *ADAMTS2* | 0.0021 | 0.0010 | 4.23E-02 | ++ | 0.00158 | 0.00251 | 0.62820 | 5.34E-01 | 0.00222 | 0.00114 | 1.94506 | 5.88E-02 |
| Synaptic density | Asyn | *SLC38A2* | 0.0000 | 0.0001 | 7.85E-01 | ++ | 0.00001 | 0.00009 | 0.12280 | 9.03E-01 | 0.00005 | 0.00016 | 0.33681 | 7.38E-01 |
| Synaptic density | Asyn | *CHGA* | 0.0001 | 0.0001 | 2.35E-01 | -+ | -0.00013 | 0.00012 | -1.07931 | 2.87E-01 | 0.00016 | 0.00008 | 2.09608 | 4.24E-02 |
| Synaptic density | Asyn | *VAT1* | -0.0003 | 0.0003 | 3.49E-01 | +- | 0.00007 | 0.00056 | 0.11778 | 9.07E-01 | -0.00044 | 0.00037 | -1.19811 | 2.38E-01 |
| Synaptic density | Asyn | *PAFAH1B3* | 0.0009 | 0.0011 | 4.31E-01 | ++ | 0.00197 | 0.00201 | 0.97914 | 3.34E-01 | 0.00039 | 0.00139 | 0.28013 | 7.81E-01 |
| Synaptic density | Asyn | *SLC6A9* | 0.0004 | 0.0006 | 4.53E-01 | ++ | 0.00024 | 0.00067 | 0.36363 | 7.18E-01 | 0.00079 | 0.00099 | 0.80008 | 4.28E-01 |
| Synaptic density | Asyn | *TMCC2* | 0.0001 | 0.0002 | 6.56E-01 | -+ | -0.00002 | 0.00023 | -0.06996 | 9.45E-01 | 0.00015 | 0.00022 | 0.68485 | 4.97E-01 |
| Synaptic density | Asyn | *ARSG* | -0.0033 | 0.0039 | 4.06E-01 | -- | -0.00075 | 0.00576 | -0.13089 | 8.97E-01 | -0.00548 | 0.00539 | -1.01514 | 3.16E-01 |
| Synaptic density | Asyn | *IL2RG* | -0.0060 | 0.0050 | 2.27E-01 | -- | -0.00957 | 0.00854 | -1.12034 | 2.69E-01 | -0.00419 | 0.00610 | -0.68576 | 4.97E-01 |
| Synaptic density | Asyn | *HSD11B2* | 0.0038 | 0.0016 | 1.58E-02 | ++ | 0.00024 | 0.00397 | 0.06135 | 9.51E-01 | 0.00439 | 0.00169 | 2.59743 | 1.31E-02 |
| Synaptic density | Asyn | *PRELP* | -0.0001 | 0.0001 | 3.45E-01 | -- | -0.00004 | 0.00029 | -0.12846 | 8.98E-01 | -0.00016 | 0.00016 | -1.01150 | 3.18E-01 |
| Synaptic density | Asyn | *FHIP1B* | 0.0002 | 0.0008 | 8.32E-01 | +- | 0.00091 | 0.00084 | 1.07385 | 2.89E-01 | -0.00453 | 0.00213 | -2.13104 | 3.93E-02 |
| Synaptic density | Asyn | *EMP3* | 0.0003 | 0.0008 | 6.80E-01 | -+ | -0.00060 | 0.00161 | -0.37079 | 7.13E-01 | 0.00063 | 0.00092 | 0.68584 | 4.97E-01 |
| Neuroinflammatory | Iba1 | *ADAMTS2* | -0.0010 | 0.0012 | 3.96E-01 | -- | -0.00153 | 0.00296 | -0.51611 | 6.08E-01 | -0.00095 | 0.00135 | -0.69821 | 4.89E-01 |
| Neuroinflammatory | Iba1 | *SLC38A2* | 0.0000 | 0.0001 | 6.94E-01 | +- | 0.00018 | 0.00010 | 1.78609 | 8.04E-02 | -0.00034 | 0.00016 | -2.12318 | 4.00E-02 |
| Neuroinflammatory | Iba1 | *CHGA* | -0.0001 | 0.0001 | 1.90E-01 | -- | -0.00017 | 0.00011 | -1.51058 | 1.37E-01 | -0.00004 | 0.00008 | -0.53893 | 5.93E-01 |
| Neuroinflammatory | Iba1 | *VAT1* | -0.0003 | 0.0003 | 3.62E-01 | +- | 0.00105 | 0.00065 | 1.61931 | 1.12E-01 | -0.00074 | 0.00037 | -1.98093 | 5.45E-02 |
| Neuroinflammatory | Iba1 | *PAFAH1B3* | -0.0009 | 0.0012 | 4.49E-01 | -- | -0.00189 | 0.00184 | -1.02608 | 3.10E-01 | -0.00022 | 0.00149 | -0.14474 | 8.86E-01 |
| Neuroinflammatory | Iba1 | *SLC6A9* | 0.0005 | 0.0006 | 3.72E-01 | +- | 0.00148 | 0.00072 | 2.04459 | 4.64E-02 | -0.00129 | 0.00099 | -1.29520 | 2.03E-01 |
| Neuroinflammatory | Iba1 | *TMCC2* | -0.0004 | 0.0002 | 3.31E-02 | +- | 0.00003 | 0.00027 | 0.10560 | 9.16E-01 | -0.00062 | 0.00022 | -2.81583 | 7.52E-03 |
| Neuroinflammatory | Iba1 | *ARSG* | 0.0085 | 0.0039 | 3.11E-02 | -+ | -0.00176 | 0.00675 | -0.26135 | 7.95E-01 | 0.01371 | 0.00483 | 2.83787 | 7.10E-03 |
| Neuroinflammatory | Iba1 | *IL2RG* | -0.0016 | 0.0045 | 7.19E-01 | -- | -0.00352 | 0.00788 | -0.44630 | 6.57E-01 | -0.00070 | 0.00543 | -0.12883 | 8.98E-01 |
| Neuroinflammatory | Iba1 | *HSD11B2* | -0.0021 | 0.0019 | 2.56E-01 | -- | -0.00424 | 0.00467 | -0.90770 | 3.69E-01 | -0.00171 | 0.00202 | -0.84405 | 4.04E-01 |
| Neuroinflammatory | Iba1 | *PRELP* | -0.0003 | 0.0001 | 2.99E-02 | -- | -0.00008 | 0.00030 | -0.27679 | 7.83E-01 | -0.00036 | 0.00015 | -2.30125 | 2.67E-02 |
| Neuroinflammatory | Iba1 | *FHIP1B* | 0.0016 | 0.0008 | 4.69E-02 | ++ | 0.00160 | 0.00089 | 1.80064 | 7.80E-02 | 0.00189 | 0.00223 | 0.84921 | 4.01E-01 |
| Neuroinflammatory | Iba1 | *EMP3* | 0.0009 | 0.0008 | 2.97E-01 | ++ | 0.00096 | 0.00189 | 0.51066 | 6.12E-01 | 0.00085 | 0.00093 | 0.91066 | 3.68E-01 |

**Supplementary Table 7:** Association analysis between 13 DEGs and neuropathological traits in each of SymAD, AsymAD and Control groups of FHS. Only P-values less than 0.05 are shown.

| Group | Gene | Traits | Beta | SE | Tvalue | Pvalue |
|---|---|---|---|---|---|---|
| SymAD | *ADAMTS2* | ICAM1 | 0.0108 | 0.0029 | 3.7779 | 5.57E-04 |
| SymAD | *ADAMTS2* | pTau231 | 0.0104 | 0.0030 | 3.4549 | 1.40E-03 |
| SymAD | *ADAMTS2* | pTau181 | 0.0086 | 0.0031 | 2.8087 | 7.90E-03 |
| SymAD | *ADAMTS2* | Ab42 | -0.0054 | 0.0026 | -2.0798 | 4.45E-02 |
| SymAD | *SLC38A2* | BraakStage | 0.0002 | 0.0001 | 2.2460 | 3.08E-02 |
| SymAD | *CHGA* | pTau231 | 0.0005 | 0.0002 | 2.4860 | 1.76E-02 |
| SymAD | *CHGA* | Ab42 | -0.0004 | 0.0002 | -2.2592 | 2.98E-02 |
| SymAD | *CHGA* | pTau396 | 0.0005 | 0.0002 | 2.2127 | 3.33E-02 |
| SymAD | *CHGA* | bFGF | -0.0005 | 0.0002 | -2.1249 | 4.03E-02 |
| SymAD | *VAT1* | pTau231 | 0.0024 | 0.0008 | 3.1886 | 2.91E-03 |
| SymAD | *VAT1* | pTau396 | 0.0024 | 0.0009 | 2.6998 | 1.05E-02 |
| SymAD | *PAFAH1B3* | BraakStage | 0.0060 | 0.0018 | 3.3304 | 1.97E-03 |
| SymAD | *PAFAH1B3* | CERAD | 0.0054 | 0.0019 | 2.8845 | 6.50E-03 |
| SymAD | *PAFAH1B3* | Arteriosclerosis | 0.0094 | 0.0035 | 2.7084 | 1.02E-02 |
| SymAD | *SLC6A9* | pTau396 | 0.0032 | 0.0009 | 3.6533 | 8.18E-04 |
| SymAD | *SLC6A9* | pTau202 | 0.0026 | 0.0010 | 2.6909 | 1.07E-02 |
| SymAD | *SLC6A9* | pTau181 | 0.0021 | 0.0009 | 2.4286 | 2.01E-02 |
| SymAD | *TMCC2* | pTau396 | 0.0011 | 0.0003 | 3.6985 | 7.19E-04 |
| SymAD | *TMCC2* | pTau202 | 0.0010 | 0.0003 | 3.0049 | 4.82E-03 |
| SymAD | *ARSG* | AT8 | -0.0116 | 0.0056 | -2.0798 | 4.47E-02 |
| SymAD | *IL2RG* | pTau202 | -0.0237 | 0.0111 | -2.1369 | 3.95E-02 |
| SymAD | *HSD11B2* | ICAM1 | 0.0156 | 0.0052 | 3.0057 | 4.74E-03 |
| SymAD | *HSD11B2* | pTau231 | 0.0135 | 0.0055 | 2.4654 | 1.84E-02 |
| SymAD | *HSD11B2* | pTau181 | 0.0124 | 0.0054 | 2.2927 | 2.76E-02 |
| SymAD | *HSD11B2* | Flt1 | 0.0107 | 0.0048 | 2.2330 | 3.17E-02 |
| SymAD | *HSD11B2* | Atherosclerosis | 0.0143 | 0.0066 | 2.1834 | 3.79E-02 |
| SymAD | *PRELP* | CRP | -0.0007 | 0.0003 | -2.2850 | 2.81E-02 |
| SymAD | *FHIP1B* | Arteriosclerosis | -0.0097 | 0.0034 | -2.8275 | 7.53E-03 |
| SymAD | *FHIP1B* | Iba1 | -0.0071 | 0.0027 | -2.5935 | 1.37E-02 |
| AsymAD | *SLC38A2* | Flt1 | -0.0006 | 0.0002 | -2.7446 | 2.87E-02 |
| AsymAD | *SLC38A2* | ord_CAA | 0.0006 | 0.0002 | 2.6371 | 3.36E-02 |
| AsymAD | *CHGA* | pTau396 | -0.0010 | 0.0003 | -3.8322 | 6.44E-03 |
| AsymAD | *VAT1* | C4B | 0.0096 | 0.0029 | 3.3464 | 1.55E-02 |
| AsymAD | *VAT1* | CRP | 0.0056 | 0.0021 | 2.6028 | 3.53E-02 |
| AsymAD | *VAT1* | Iba1 | 0.0047 | 0.0018 | 2.5785 | 3.66E-02 |
| AsymAD | *PAFAH1B3* | pTau396 | -0.0048 | 0.0014 | -3.3923 | 1.16E-02 |
| AsymAD | *SLC6A9* | CAA | 0.0093 | 0.0025 | 3.7227 | 7.43E-03 |
| AsymAD | *TMCC2* | C4A | 0.0011 | 0.0004 | 3.0420 | 2.27E-02 |
| AsymAD | *IL2RG* | Ab42 | -0.0872 | 0.0273 | -3.1917 | 1.52E-02 |
| AsymAD | *IL2RG* | AB4G8 | 0.0901 | 0.0364 | 2.4777 | 4.24E-02 |
| AsymAD | *IL2RG* | Vegf.C | 0.0948 | 0.0395 | 2.4019 | 4.73E-02 |
| AsymAD | *HSD11B2* | Vegf.C | -0.0268 | 0.0102 | -2.6360 | 3.36E-02 |
| AsymAD | *PRELP* | PSD.95 | -0.0033 | 0.0008 | -4.0158 | 5.09E-03 |
| AsymAD | *FHIP1B* | C4A | -0.0107 | 0.0040 | -2.6784 | 3.66E-02 |
| AsymAD | *EMP3* | Vegf.D | 0.0076 | 0.0031 | 2.4871 | 4.18E-02 |
| AsymAD | *EMP3* | pTau396 | -0.0052 | 0.0022 | -2.4024 | 4.73E-02 |
| Control | *ADAMTS2* | pTau396 | -0.0036 | 0.0014 | -2.6710 | 9.55E-03 |
| Control | *SLC38A2* | C4B | 0.0002 | 0.0001 | 2.7867 | 7.03E-03 |
| Control | *CHGA* | VCAM1 | -0.0004 | 0.0001 | -2.8304 | 6.11E-03 |
| Control | *CHGA* | Ab42 | -0.0004 | 0.0002 | -2.3929 | 1.95E-02 |
| Control | *VAT1* | pTau396 | -0.0009 | 0.0003 | -3.2786 | 1.68E-03 |
| Control | *VAT1* | Vegf.C | 0.0006 | 0.0003 | 2.0413 | 4.51E-02 |
| Control | *PAFAH1B3* | aSyn | 0.0061 | 0.0022 | 2.7849 | 7.45E-03 |
| Control | *SLC6A9* | pTau231 | 0.0020 | 0.0006 | 3.2726 | 1.70E-03 |
| Control | *SLC6A9* | SAA | 0.0022 | 0.0007 | 2.9416 | 4.46E-03 |
| Control | *SLC6A9* | aSyn | 0.0022 | 0.0009 | 2.3957 | 2.02E-02 |
| Control | *TMCC2* | VCAM1 | -0.0005 | 0.0002 | -2.1315 | 3.67E-02 |
| Control | *TMCC2* | C1q | -0.0006 | 0.0003 | -2.0588 | 4.36E-02 |
| Control | *TMCC2* | CD68 | -0.0005 | 0.0002 | -2.0549 | 4.37E-02 |
| Control | *ARSG* | pTau396 | -0.0072 | 0.0031 | -2.3585 | 2.14E-02 |
| Control | *ARSG* | VCAM1 | 0.0073 | 0.0031 | 2.3521 | 2.16E-02 |
| Control | *ARSG* | aSyn | -0.0085 | 0.0041 | -2.0508 | 4.53E-02 |
| Control | *IL2RG* | pTau231 | -0.0126 | 0.0047 | -2.6919 | 8.99E-03 |
| Control | *IL2RG* | Ab40 | -0.0122 | 0.0052 | -2.3309 | 2.27E-02 |
| Control | *IL2RG* | CD68 | 0.0112 | 0.0052 | 2.1642 | 3.40E-02 |
| Control | *IL2RG* | pTau396 | -0.0102 | 0.0050 | -2.0423 | 4.52E-02 |
| Control | *PRELP* | C4B | 0.0007 | 0.0002 | 3.7906 | 3.39E-04 |
| Control | *PRELP* | Flt1 | -0.0006 | 0.0002 | -3.0829 | 2.98E-03 |
| Control | *PRELP* | PIGF | -0.0006 | 0.0002 | -2.8746 | 5.42E-03 |
| Control | *PRELP* | Tie2 | -0.0006 | 0.0002 | -2.7642 | 7.34E-03 |
| Control | *PRELP* | pTau231 | -0.0005 | 0.0002 | -2.6987 | 8.83E-03 |
| Control | *PRELP* | ICAM1 | -0.0004 | 0.0002 | -2.3005 | 2.45E-02 |
| Control | *PRELP* | pTau181 | -0.0004 | 0.0002 | -2.1967 | 3.15E-02 |
| Control | *PRELP* | Iba1 | -0.0004 | 0.0002 | -2.1219 | 3.75E-02 |
| Control | *FHIP1B* | C4B | -0.0056 | 0.0020 | -2.8241 | 6.34E-03 |
| Control | *FHIP1B* | CD68 | 0.0046 | 0.0017 | 2.6569 | 9.82E-03 |
| Control | *FHIP1B* | Iba1 | 0.0041 | 0.0018 | 2.2668 | 2.66E-02 |
| Control | *FHIP1B* | Atherosclerosis | -0.0046 | 0.0021 | -2.2241 | 3.04E-02 |
| Control | *FHIP1B* | PIGF | 0.0042 | 0.0021 | 2.0603 | 4.33E-02 |
| Control | *EMP3* | Flt1 | 0.0027 | 0.0007 | 3.6885 | 4.54E-04 |
| Control | *EMP3* | pTau396 | -0.0021 | 0.0006 | -3.2537 | 1.81E-03 |
| Control | *EMP3* | PIGF | 0.0020 | 0.0008 | 2.5657 | 1.25E-02 |

**Supplementary Table 8:** 52 DEGs between SymAD and AsymAD in Bellenguez's AD GWAS study. Two genes (*IL2RG* and *SMPX*) in chromosome X were excluded.

| Gene | Nearby_Gene | Region | SNP | CHR | BP | P-value |
|---|---|---|---|---|---|---|
| *ADAMTS2* | *ADAMTS2* | intronic | rs549839389 | 5 | 179266217 | 3.34E-04 |
| *S100A4* | *S100A2(dist=12141),S100A16(dist=28898)* | intergenic | rs74660920 | 1 | 153577985 | 4.67E-03 |
| *NRIP2* | *ITFG2* | intronic | rs117701901 | 12 | 2814087 | 1.16E-03 |
| *SCGN* | *SCGN* | intronic | rs144820707 | 6 | 25690175 | 4.15E-03 |
| *SLC38A2* | *SLC38A1(dist=82551),SLC38A2(dist=6594)* | intergenic | rs150814412 | 12 | 46351594 | 3.50E-03 |
| *ALDH1A1* | *ALDH1A1(dist=105878),ANXA1(dist=92934)* | intergenic | rs555747583 | 9 | 73058931 | 2.46E-04 |
| *CHGA* | *ITPK1(NM_014216:c.*2488A>G,NM_001142593:c.*2488A>G)* | UTR3 | rs146378637 | 14 | 92939073 | 2.17E-03 |
| *CEP83* | *PLXNC1* | intronic | rs832505 | 12 | 94278959 | 2.12E-04 |
| *VAT1* | *RUNDC1(dist=442)* | downstream | rs139850417 | 17 | 42995584 | 1.85E-03 |
| *PAFAH1B3* | *TMEM145* | intronic | rs779361907 | 19 | 42323038 | 8.05E-04 |
| *POLD1* | *MYBPC2* | intronic | rs7258429 | 19 | 50435297 | 5.79E-04 |
| *IVD* | *IVD* | intronic | rs74834275 | 15 | 40427879 | 6.44E-03 |
| *ARG2* | *ARG2* | intronic | rs561778116 | 14 | 67643110 | 7.23E-04 |
| *SLC6A9* | *ATP6V0B* | intronic | rs72890653 | 1 | 43975990 | 3.71E-03 |
| *TMCC2* | *DSTYK* | intronic | rs34119249 | 1 | 205200346 | 8.07E-04 |
| *CLIP2* | *RFC2(dist=22996),CLIP2(dist=11953)* | intergenic | rs7798046 | 7 | 74277454 | 1.14E-04 |
| *ARSG* | *ARSG* | intronic | rs147304255 | 17 | 68398540 | 1.34E-03 |
| *QDPR* | *SNORA75B(dist=110691),QDPR(dist=54820)* | intergenic | rs756228995 | 4 | 17431573 | 1.35E-03 |
| *NSMCE1* | *NSMCE1* | intronic | rs191181316 | 16 | 27242690 | 1.92E-03 |
| *SCG3* | *SCG3(NM_001165257:c.*1451T>C,NM_013243:c.*1451T>C)* | UTR3 | rs184552675 | 15 | 51720977 | 2.38E-03 |
| *CXCR4* | *CXCR4(dist=8762),THSD7B(dist=638628)* | intergenic | rs545606749 | 2 | 136126917 | 4.91E-03 |
| *RABGGTA* | *DHRS1* | intronic | rs10146996 | 14 | 24294980 | 2.19E-03 |
| *RAB11FIP5* | *RAB11FIP5(dist=14088),NOTO(dist=75538)* | intergenic | rs562226656 | 2 | 73127036 | 9.80E-04 |
| *HSD11B2* | *ATP6V0D1* | intronic | rs148869109 | 16 | 67455321 | 7.91E-04 |
| *SAP30L* | *HAND1(dist=7380),MIR3141(dist=110405)* | intergenic | rs182243 | 5 | 154485607 | 1.52E-04 |
| *ZBBX* | *LINC01327(dist=17046),SERPINI2(dist=16224)* | intergenic | rs149657876 | 3 | 167425565 | 6.17E-03 |
| *USP31* | *USP31* | intronic | rs750344696 | 16 | 23114001 | 3.74E-03 |
| *SLC43A3* | *SLC43A3(dist=27528),RTN4RL2(dist=5420)* | intergenic | rs187168722 | 11 | 57455108 | 1.17E-03 |
| *LRATD2* | *LRATD2(dist=2323),PCAT1(dist=452353)* | intergenic | rs147940485 | 8 | 126560801 | 4.89E-03 |
| *MTMR10* | *MTMR10* | intronic | rs180784786 | 15 | 30946184 | 9.82E-04 |
| *PRDM8* | *PRDM8(dist=9426),FGF5(dist=52834)* | intergenic | rs62300378 | 4 | 80213754 | 8.18E-04 |
| *DYNLRB1* | *DYNLRB1* | intronic | rs562493105 | 20 | 34528027 | 2.95E-05 |
| *TNFRSF18* | *B3GALT6(dist=680)* | downstream | rs12096608 | 1 | 1235720 | 1.18E-02 |
| *MON1B* | *MIR4719(dist=301974),MON1B(dist=20197)* | intergenic | rs761640542 | 16 | 77170993 | 5.01E-04 |
| *PEPD* | *PEPD* | intronic | rs116601945 | 19 | 33388625 | 3.44E-04 |
| *WBP2NL* | *SEPTIN3* | intronic | rs2269665 | 22 | 41981874 | 4.66E-04 |
| *ELP2* | *SLC39A6(NM_012319:c.*588A>G)* | UTR3 | rs909774216 | 18 | 36109005 | 4.20E-03 |
| *GEM* | *GEM(dist=25987),RAD54B(dist=83654)* | intergenic | rs187174139 | 8 | 94288306 | 2.18E-03 |
| *PRELP* | *PRELP* | intronic | rs79034827 | 1 | 203480711 | 1.02E-02 |
| *FHIP1B* | *CNGA4* | exonic | rs186291453 | 11 | 6240112 | 9.34E-06 |
| *MARCHF8* | *MARCHF8* | intronic | rs769718720 | 10 | 45565871 | 4.79E-04 |
| *MED29* | *PLEKHG2(NM_001351693:c.-1174T>C,NM_022835:c.-1174T>C)* | UTR5 | rs114784999 | 19 | 39412913 | 5.05E-03 |
| *EMP3* | *CCDC114* | intronic | rs140531044 | 19 | 48310574 | 1.53E-03 |
| *ANP32E* | *ANP32E(dist=16480),CA14(dist=5182)* | intergenic | rs181209704 | 1 | 150252592 | 1.13E-03 |
| *CNKSR3* | *IPCEF1(dist=11250),CNKSR3(dist=19499)* | intergenic | rs2473550 | 6 | 154368016 | 8.28E-04 |
| *PRR13* | *SP1(dist=1433),AMHR2(dist=5980)* | intergenic | rs574646923 | 12 | 53417875 | 2.12E-03 |
| *PPM1D* | *PPM1D* | intronic | rs141144548 | 17 | 60616074 | 5.25E-03 |
| *MYOT* | *MYOT* | intronic | rs185757192 | 5 | 137884799 | 2.27E-03 |
| *CHN2* | *CHN2* | intronic | rs62459612 | 7 | 29343267 | 3.02E-04 |
| *STAG1* | *STAG1* | intronic | rs139270227 | 3 | 136716306 | 1.90E-06 |
